# Youth Correlates of Genetic Liability to Substance Use Disorders

**DOI:** 10.1101/2025.11.22.25340798

**Authors:** Sarah E. Paul, Aaron J. Gorelik, Nicole R. Karcher, Alex P. Miller, David A.A. Baranger, Emma C. Johnson, Kimberly H. LeBlanc, Gayathri Dowling, Deanna M. Barch, Alexander S. Hatoum, Arpana Agrawal, Ryan Bogdan

## Abstract

Substance use disorders (SUDs) are moderately-highly heritable and develop in stages that typically begin in adolescence/young adulthood. It remains unclear whether SUD correlates reflect predispositional risk and/or arise as consequences of exposure. Our phenome-wide association study (PheWAS) of polygenic risk for SUDs (i.e., Problematic Alcohol Use [PAU], Tobacco Use Disorder [TUD], Cannabis Use Disorder [CUD], Opioid Use Disorder [OUD], and General Addiction Risk Factor [ADDrf]) in 1,584-5,556 8-13-year-old children enrolled in the Adolescent Brain Cognitive Development (ABCD) Study identified 309 phenotypic associations, particularly among mental health, sleep, screen time, and caffeine consumption phenotypes. Most associations were observed in substance naïve participants. Neuroimaging phenotypes (e.g., resting state functional connectivity) partially accounted for associations between SUD PRS and screen time phenotypes. Genetic liability to SUDs is expressed during childhood as potentially modifiable risk factors that could be targeted to attenuate its expression in adolescence and adulthood.

## Main

Substance use disorders (SUDs) are common and heritable (50-60%)^1,2^ conditions that significantly adversely impact individuals, families, and society^3–5^. Typically, the first serious symptoms of SUDs arise in late adolescence and/or early adulthood. Identification of the compendium of early markers of genetic susceptibility to SUDs may highlight mechanisms through which predispositional risk manifests that could be targeted to reduce risk.

Here, to identify correlates of genetic liability to SUDs in childhood, we conduct Phenome-wide Association Studies (PheWASs) of polygenic scores (PRS) for Problematic Alcohol Use (PAU), Tobacco Use Disorder (TUD), Cannabis Use Disorder (CUD), Opioid Use Disorder (OUD), and a general Addiction Risk Factor (ADDrf) in individuals genetically similar to African and European reference populations in the Adolescent Brain Cognitive Development Study^SM^ (ABCD Study®) at ages 8-11 (baseline; n=1,584, 5,556, respectively) and 10-13 (follow-up; n=1,273, 5,048). We test associations between each PRS and up to 1,697 psychosocial and health-related phenotypes and 421 imaging-derived phenotypes. Follow-up analyses exclude participants who had initiated substance use to rule out potential confounding by concurrent substance use. Ultimately, phenotypes identified as correlates of genetic liability to SUDs during childhood, prior to SUD expression, and in many cases, substance exposures, may reflect putative risk factors through which genetic liability is expressed. These phenotypes could serve as accessible, malleable targets for family-based education/intervention to improve the health of youth at genetic risk for SUDs.

## Results

### Non-imaging

#### European-like ancestry

See **Table 1** for sample descriptive information.

**Table 1.**
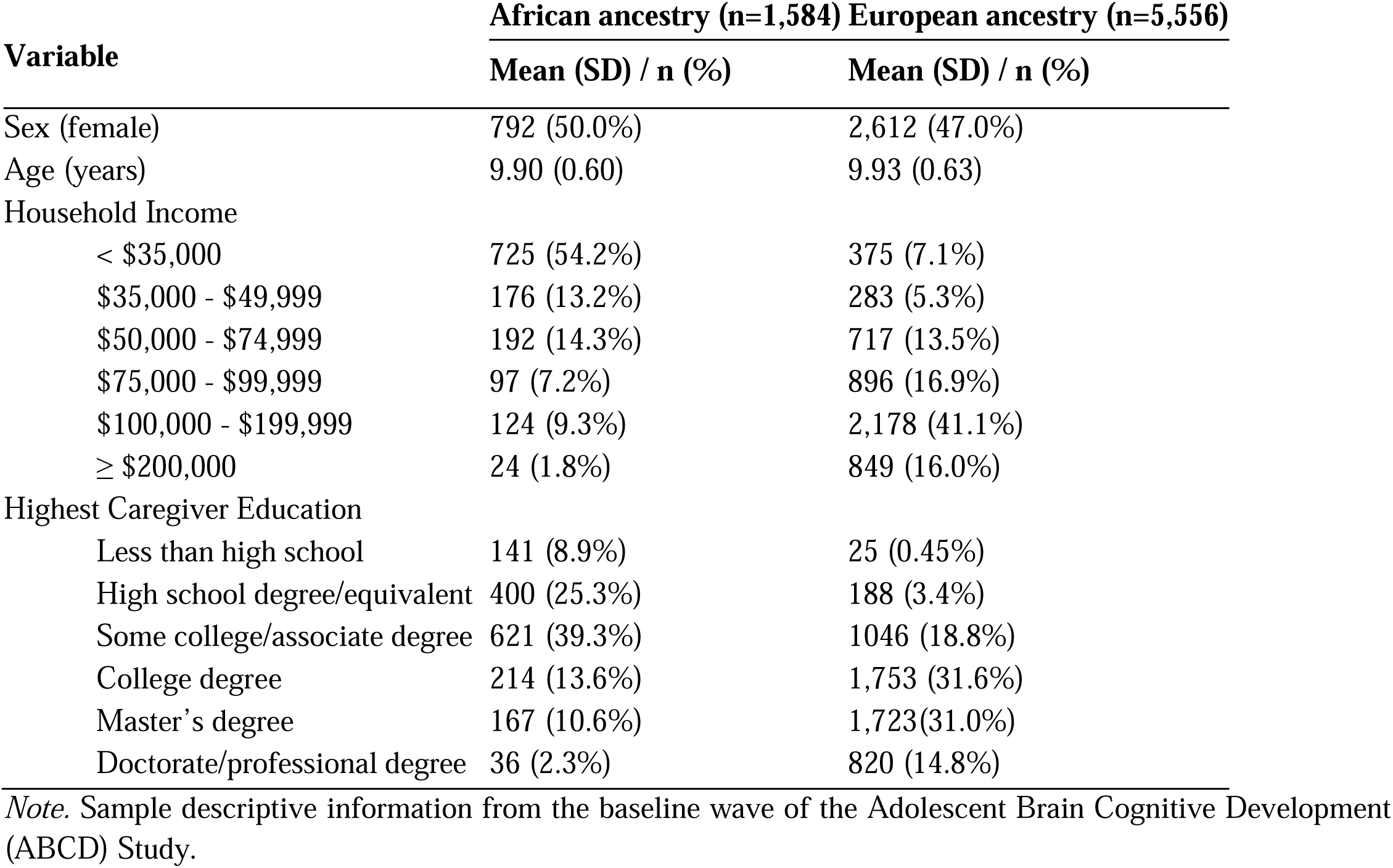
Sample descriptive statistics.

#### Broad Patterns

##### Number of Significant Associations

Each substance use disorder PRS (SUD_PRS;_ PAU, TUD, CUD, OUD, ADDrf; **Table 2**) was associated with 10-71 baseline (0.79-5.59%) and 19-136 follow-up (1.12-8.01%) phenotypes following Bonferroni correction (*p_baseline_*<3.93e-05, *p_follow-up_*<2.95e-05), with their average absolute standardized betas and ORs ranging from 0.049-0.074 (avg=0.061) and 1.25-1.40 (avg=1.32), respectively (**Figures 1-2; Supplemental Tables 9-10; Extended Data Figures 1-2)**. ***Associations by Domain.*** Across time points but varying by PRS, the screentime/media use, cognition, substance use and perceptions, and school-related domains had the greatest percentage of significantly associated phenotypes (up to 50%; **Table 3**; **Figure 2; Supplemental Note; Supplemental Tables 9-10**).

**Figure 1.**
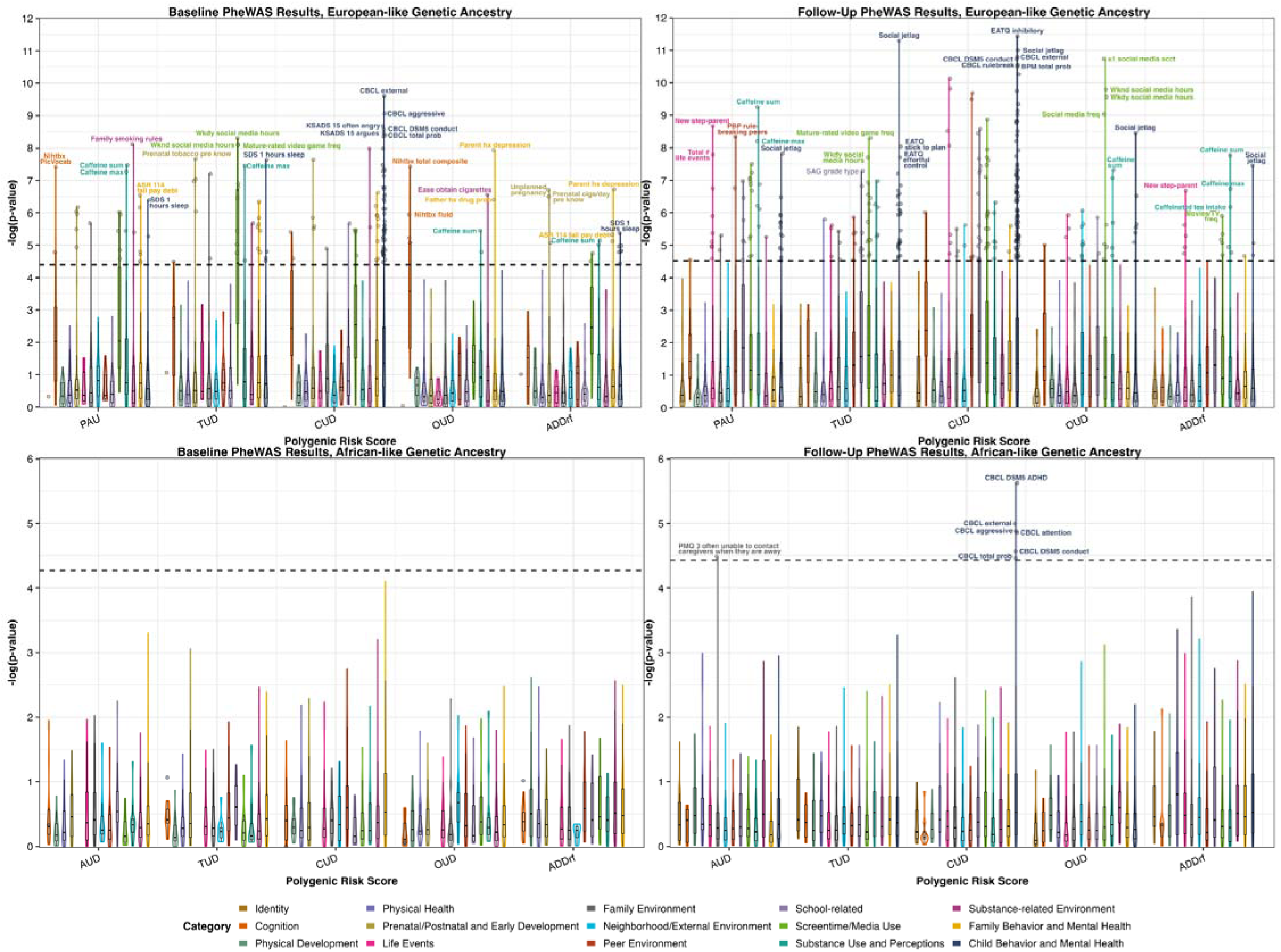
*Note.* Manhattan and violin plots depicting the number, distribution, and types of phenotypes, within each category, associated with each of the 5 examined PRS. The x-axes are the 5 PRS, the y-axes are the negative log of the p-value of association (higher value indicates stronger association), and color indicates phenotypic Category. The dashed horizontal lines denote the Bonferroni-significant threshold (i.e., 0.05/number of phenotypes). The violin plots represent the distribution of -log(p-values) within each Category, separately for each PRS. Boxplots within each violin plot show the median -log(p-value of association), and the boxes encompass the middle 50% of associations. PAU = Problematic Alcohol Use; TUD = Tobacco Use Disorder; CUD = Cannabis Use Disorder; OUD = Opioid Use Disorder; ADDrf = Addiction Risk Factor; AUD = Alcohol Use Disorder.

**Figure 2.**
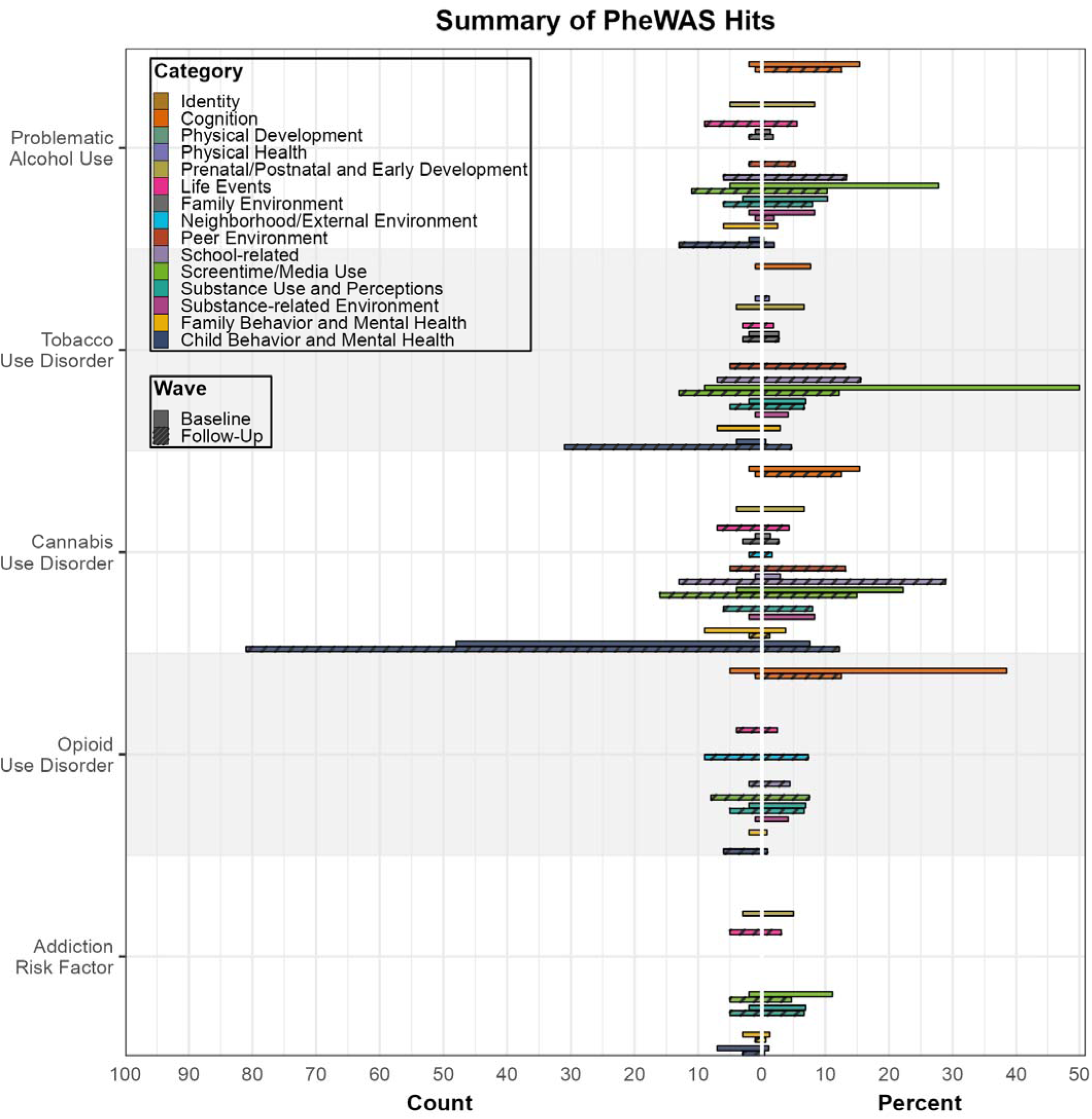
Note. Summary of the number (“Count,” left) and percent (right) of associations observed between each PRS (y-axis) and phenotypes in each category, for European-like genetic ancestry. Colors denotes different categories, and solid and dashed bars represent Baseline and Two-Year Follow-Up associations, respectively. Percent reflects the percent of phenotypes in a category associat d with a particular PRS of the total number of phenotypes in that category.

**Table 2.** Discovery GWAS Used to Generate PRS.

| **GWAS Phenotype** | **Ancestry and n** |
| --- | --- |
| Alcohol Use Disorder(AUD)/Problematic Alcohol Use (PAU)<sup>3a</sup> | AUD: African n=122,571 supplemented with European n=739,895<br>PAU: European n=903,147 |
| Tobacco Use Disorder (TUD)<sup>35</sup> | African n=114,420<sup>b</sup><br>European n=739,895 |
| Cannabis Use Disorder (CUD)<sup>36</sup> | African n=123,208<sup>b</sup>; European n=886,025 |
| Opioid Use Disorder (OUD)<sup>34</sup> | African n=84,877<sup>b</sup>; European n=554,186 |
| Addiction Risk Factor (ADDrf<sup>40</sup>) | African n=92,630<sup>b</sup>; European n=1,025,550 |

*Note.* <sup>a</sup>The cross-ancestry genetic-effect correlation between PAU and AUD is 0.71<sup>3</sup>. <sup>b</sup>African ancestry samples were supplemented with the European ancestry sample in PRS-CSx.

**Table 3.**
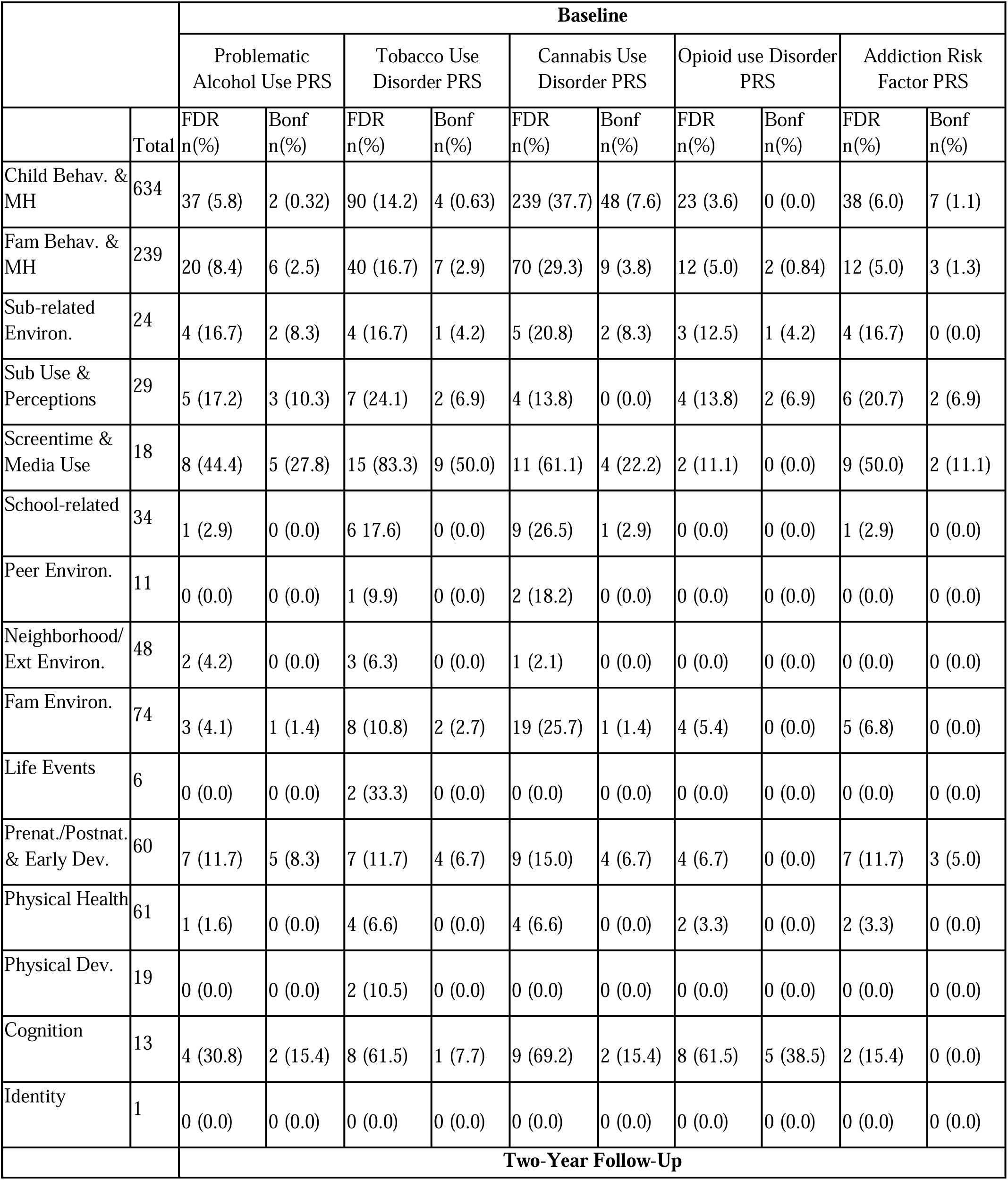
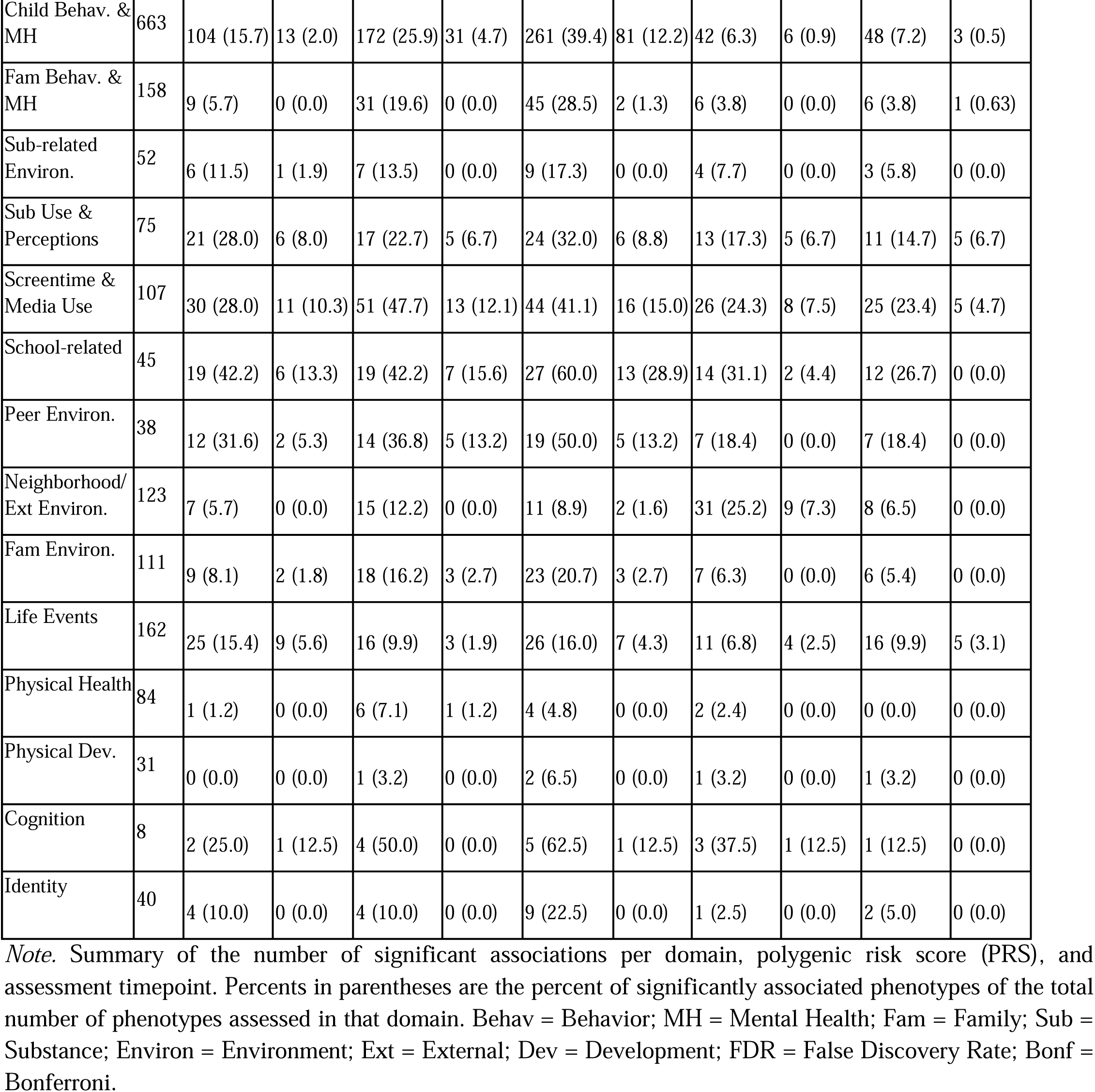
Summary of associations by phenotype domain.

### Child Behavior and Mental Health

SUD_PRS_--especially CUD_PRS_ and TUD_PRS_--exhibited multiple associations with mental health phenotypes (0.027≤|B|≤0.069; 1.16≤OR^1^ 1.59), particularly those related to externalizing behavior. Notably, CUD_PRS_ was associated with a significantly greater proportion of child behavior and psychopathology--particularly externalizing--phenotypes compared to all other PRS (baseline χ2(1)≥40.4, *p*≤3.50e-08; follow-up χ2(1)≥23.4, *p*≤1.31e-06). Several non-externalizing associations also emerged, including greater psychotic-like experiences (PAU_PRS_, TUD_PRS_, and CUD_PRS_), youth- or teacher-reported total behavior problems (all except ADDrf_PRS_), and greater sleep problems (e.g., fewer hours, total sleep disturbances, and social jetlag [i.e., discrepancy between weekday and weekend sleep times]).

### Screentime/Media Use

Each SUD_PRS_ was positively associated with frequency and time spent using social media. All SUD_PRS_, except OUD_PRS_, were associated with weekend screen time and R-rated movie frequency, with additional SUD-specific associations observed (e.g., higher frequency of playing mature video games, as well as weekday screen weekend texting, and weekday TV/movie time (0.038≤|B|≤0.080; 1.15≤OR≤1.26).

### Cognition

OUD_PRS_ was negatively associated with more phenotypes related to cognitive performance (e.g., NIH Toolbox total composite, fluid intelligence,) at baseline (n=5) than any other PRS (n≤2) (across waves and PRS, 0.054≤|B|≤0.074). Cognition phenotypes represented 50% of significant baseline phenotypic associations with OUD_PRS_.

### Substance Use and Perceptions

All SUD_PRS_ were associated with caffeine intake (e.g., maximum intake, use summary score, and/or energy drink). Associations also emerged for less perceived harm for regular smokeless tobacco use (all PRS) and more positive cannabis use expectancies (CUD_PRS_; 0.027≤|B|≤0.083; 1.16≤OR≤1.18).

### School-related

Only CUD_PRS_ was associated lower school performance at baseline, and all PRS except ADDrf_PRS_ were linked with lower school performance, increased likelihood of detention, and/or school disengagement, phenotypes at follow-up (0.038≤|B|≤0.088; 1.21≤OR≤1.46).

### Prenatal/Postnatal and Early Development

With the exception of OUD_PRS_, all SUD_PRS_ were associated with unplanned pregnancy and prenatal tobacco exposure; PAU_PRS_ and CUD_PRS_ were additionally associated with prenatal cannabis exposure (0.023≤|B|≤0.035; 1.20≤OR≤1.65).

### Family Behavior and Mental Health

All SUD_PRS_ were positively associated with paternal history of drug problems and parental history of depression. Additional positive associations emerged, including with current caregiver externalizing-related and somatic problems (TUD_PRS_ and CUD_PRS_; 0.025≤|B|≤0.059; 1.25≤OR≤1.46).

### Substance-related Environment

PAU_PRS_, TUD_PRS_, CUD_PRS_, and OUD_PRS_ were associated with the presence of differential cigarette use rules among family members, greater cigarette accessibility, and/or peer tobacco use (0.029≤|B|≤0.085).

### Neighborhood/External Environment

At the follow-up wave, CUD_PRS_ was positively associated with the experience of sexual orientation- and weight-related discrimination (1.36≤OR≤1.45); OUD_PRS_ was positively coupled with several neighborhood adversity metrics (e.g., lower high-skill employment and child opportunity; higher number of individuals with disability; 0.029≤|B|≤0.11).

### Life Events

All PRS were positively associated with stressful life event exposure measured at follow-up; all PRS except OUD_PRS_ were also coupled with greater impact of these events (0.059≤|B|≤0.094; 1.18≤OR≤1.41).

### Family Environment

SUD_PRS_ were associated with lower parent/caregiver educational attainment (TUD_PRS_, CUD_PRS)_, a parent/caregiver partner not being a biological parent (PAU_PRS_, TUD_PRS)_, caregiver-child conflict (TUD_PRS_, CUD_PRS_), and less frequent family dinners (PAU; 0.043≤|B|≤0.067; 1.18≤OR≤1.50).

### Peer Environment

At the follow-up wave, PAU_PRS_, TUD_PRS_, and CUD_PRS_ were positively associated with involvement with peers who break rules and have been suspended; TUD_PRS_ and CUD_PRS_ were associated with experiencing peer victimization (OR=1.21; 0.040≤|B|≤0.092).

### Physical Health

TUD_PRS_ was negatively associated with the number of extracurricular (largely sports) activities at follow-up (B=-0.066).

### Physical Development and Identity

No significant associations were observed with any phenotypes within the Physical Development or Identity domains (B≤0.066, OR≤1.09, p≥3.72e-05, p_Bonferroni_≥0.063).

### Secondary analyses

#### Substance-naive

A majority (76.9%-100%) of Bonferroni-significant associations in the full sample were Bonferroni-significant in the substance-naive subsample (**Supplemental Tables 11-12**). ***SUD_PRS_ Specificity.*** CUD_PRS_, OUD_PRS_, and TUD_PRS_, but not PAU_PRS_ or ADDrf_PRS_, exhibited unique associations when all PRS were entered simultaneously (**Supplemental Note; Supplemental Tables S13-14**). ***Phenotypic Pruning.*** Pruning intercorrelated phenotypes resulted in a domain non-specific monotonic reduction of significant associations (range=0-66.7%; **Supplemental Tables 9-10; Extended Data Figure 3; Supplemental Note**). ***Phenotypic Change.*** No SUD_PRS_×Time interactions emerged suggesting stability of findings from Baseline to follow-up (**Supplement Note; Supplemental Table 15**). ***Sex Moderation.*** Four SUD_PRS_×sex interactions were observed (**Supplemental Note; Supplemental Tables 16-17; Extended Data Figure 4**). ***Within-family.*** Few phenotypes were associated with within-family deviations in PRS following Bonferroni correction (i.e., PAU_PRS_: increased caffeinated soda consumption; TUD_PRS_: increased caregiver restlessness, greater mood instability; CUD_PRS_: increased externalizing behavior **Supplemental Tables 18-19; Extended Data Figure 5**). ***Accounting for baseline externalizing***. Over half (58.2%-100%) of follow-up phenotypes that were previously Bonferroni-significant remained associated with SUD_PRS_ after covarying for baseline CBCL externalizing scores (**Supplemental Table 20**). These notably included measures related to caffeine intake, screentime, and sleep disturbance.

### African-like ancestry

See **Table 1** for sample descriptive information.

Seven follow-up (and no baseline) phenotypes, primarily externalizing-related behaviors, were associated with SUD_PRS_ (**Figure 1c, d; Extended Data Figures 6-7; Supplemental Tables 21-22**). All associations were significantly correlated with associations observed in EUR-like ancestry for AUD/PAU_PRS_, TUD_PRS_, CUD_PRS_, and ADDrf_PRS_ (*r=*0.089-0.39, *p*≤1.45e-04). Significant associations remained significant in the substance-naive subsample (**Supplemental Tables 23-24**). Secondary analyses revealed: 1) unique associations between CUD_PRS_ and externalizing behaviors (**Supplemental Tables 25-26**), 2) no significant SUD_PRS_×Time or SUD_PRS_×Sex interactions (**Supplemental Tables 27-29**), and e) that all CUD_PRS_ associations remained significant in within-family analyses (**Supplemental Note; Supplemental Tables 30-31**).

### Brain Imaging Results

Results were only significant in the EUR-like subsample. SUD_PRS_ were linked to RSFC (greater sensorimotor hand and sensorimotor mouth network connectivity [PAU_PRS_, CUD_PRS_]; frontoparietal and sensorimotor hand networks [CUD_PRS_]) and DWI (lower fractional anisotropy in the right corticospinal/pyramidal [TUD_PRS_], superior corticostriate [TUD_PRS_], and left superior corticostriate-parietal cortex [TUD_PRS_] tracts) metrics. These associations remained in the substance-naive subsample (**Supplement; Supplemental Tables 32-33**. ***Indirect effects results.*** All RSFC and DWI metrics significantly, indirectly linked SUD_PRS_ to follow-up phenotypes (**Supplemental Note; Extended Data Figures 8-10; Supplemental Tables 34-35**).

### Cross-Ancestry Meta-Analysis

Between 5.71 and 64.3% of Bonferroni-significant associations in EUR were similarly significant in the cross-ancestry meta-analysis. Many associations that did not survive correction in the meta-analysis exhibited heterogeneous effects across ancestry groups (**Supplemental Note; Supplemental Tables 36-37**).

## Discussion

Genetic liability to substance use disorders (i.e., PAU_PRS_, TUD_PRS_, CUD_PRS_, OUD_PRS_, ADDrf_PRS_) was strongly and most frequently associated with elevated externalizing behavior in both the EUR- and AFR-like individuals, and with screentime/media use, caffeine intake, sleep problems, family history of substance use and depression, and prenatal factors (e.g., unplanned pregnancy, prenatal substance exposure) among EUR in our PheWAS of 8-13-year-old children. Most associations persisted among substance-naive youth and thus are likely to reflect early markers of genetic liability to SUD, rather than—or in addition to—causal consequences of substance use. Collectively, this characterization of environmental, behavioral, and neural correlates of genetic liability to SUDs during childhood prior to onset of substance involvement highlights putative mechanisms through which genetic liability to substance involvement may manifest (see^6,7^ for related work in adults). Certain potentially modifiable correlates, such as screentime/media, caffeine use, and sleep should be investigated as candidates for preventative policy and intervention seeking to attenuate SUD risk.

SUD_PRS_ were strongly associated with childhood externalizing (e.g., conduct problems, aggression, inattention). Indeed, the few significant associations in AFR almost entirely comprised externalizing and related (e.g., impulsivity, school performance) phenotypes, several of which also exhibited significant cross-ancestry consistency unlike other phenotypes that were specific to EUR. These externalizing associations may overlapping genetic architecture^8^ and heterotypic continuity wherein early externalizing behaviors (e.g., ADHD, oppositional defiant disorder symptoms) represent an earlier manifestation of broad externalizing predisposition, which later includes SUDs^9^.

Caffeine intake, screentime/media use, and sleep problems—which were particularly robustly associated with SUD_PRS_—have been previously genetically and/or phenotypically correlated with externalizing problems^10–15^. However, our externalizing-adjusted PheWAS revealed that SUD_PRS_ associations with follow-up caffeine intake, screentime, and sleep disturbance were partially independent. Whether these robust residual associations are due to pleiotropy that is exclusive to SUD genetic liability, predisposition toward emotion dysregulation distinct from externalizing, and/or to other explanations is worth exploring, particularly in light of recent work in ABCD documenting prospective associations between problematic screen use and substance initiation^16^. For instance, gene-environment correlation between genetic liability to SUDs and a lack of parental supervision could contribute to excessive engagement with media and mature content viewership. In addition, youth at heightened genetic liability to SUDs were less likely to report reading for pleasure. Future studies could thus examine the extent to which the deleterious associations of media engagement could be mitigated by participation in prosocial (e.g., sporting activities, hobby clubs) and scholastic (e.g., reading, art) engagement^17^.

Reflecting the ubiquity of gene-environment correlation, genetic liability to SUDs was associated with prenatal factors (e.g., tobacco exposure, unplanned pregnancy), having rule breaking peers, experiencing victimization from and aggressing upon peers as well as experiencing stressful life events, living in a disadvantaged neighborhood and having increased ease of access to cigarettes. Particularly during childhood, genetic liability is intertwined with passive exposure to environments shaped by those genetic susceptibilities, exacerbating their dual impact^18^. These early, risky, and genotype-related environments have been linked to later substance use^19^ and variability in brain development^20^ that might hinder the emergence of the cognitive restraint necessary to deter progression to SUD, making them ideal targets for public health interventions. These associations also amplify the need to account for gene-environment correlation in research on genetic and environmental factors. To some degree, our follow-up analyses within pairs of twins and siblings evaluated this direct contribution of genetic susceptibility beyond environments experienced similarly by siblings. We found no evidence for within-family associations, although the reduced sample size combined with constrained PRS variability may have reduced statistical power. Nonetheless, given the predominance of familial environment in the etiology of initial stages of substance use^21^, many of our associations may reflect indirect pathways, from genotype to behavior, via shaping of the childhood environment.

SUD_PRS_ was robustly linked to sleep problems and less nightly sleep. Beyond the well-established acute effects of psychoactive substances and withdrawal on sleep^22,23^, growing evidence highlights a shared genetic liability underlying both sleep disruptions and substance-related phenotypes^24^, including traits like evening chronotype^25(p201)^, consistent with our social jetlag findings^26^. Whether these findings reflect yet another aspect of behavioral dysregulation, are consequent to the anti-somnolent effects of caffeine intake and protracted exposure to screens, and/or are attributable to loci with pleiotropic effects on SUD and sleep, our results suggest that youth with genetic susceptibility to SUDs should be monitored for sleep difficulties and their resultant impact on health.

Consistent with small, difficult-to-detect brain-behavior associations^27^, few statistically significant associations between SUD_PRS_ and imaging metrics emerged. However, there were two notable findings. *First*, both PAU_PRS_ and CUD_PRS_ were associated with increased resting-state functional connectivity (RSFC) between the sensorimotor hand and sensorimotor mouth networks, and CUD_PRS_ was additionally linked with increased RSFC between the frontoparietal and sensorimotor hand networks. These neural metrics indirectly linked these SUD_PRS_ to screen time behavior. A recent meta-analysis found some evidence that increased sensorimotor cortex connectivity is associated with SUD diagnosis but also with less craving, better SUD treatment response, and reduced consumption among those with SUD^28^, and another study found positive associations with the presence of gaming disorder^29^, suggesting a potential shared pathway for multiple “addictive” behaviors. *Second*, TUD_PRS_ was associated with reduced white matter integrity in the superior cortical-striatal, corticospinal/pyramidal, and cortical-striate-parietal tracts, tracts connecting regions frequently implicated in theoretical and clinical studies of addiction^30,31^, and these brain metrics partially mediated PRS association with screentime behavior. As findings were evident in substance-naive youth, they raise the intriguing possibility of preexisting brain variability associated with SUD genetic susceptibility^31^.

Lastly, while there is compelling support for genetic commonality across SUDs, captured here by ADDrf, some SUD-specific distinctions are noteworthy. CUD_PRS_ was associated with significantly more mental health phenotypes, which may be due to its high loading with other externalizing constructs^8,32^ or due to the nature of electronic health record diagnoses, which for CUD, likely reflect complicating additional psychopathology^33^. On the other hand, OUD_PRS_ was associated with fewer phenotypes overall, possibly due to its smaller size, but was uniquely associated with lower global cognitive performance and greater environmental adversity, converging with prior genetic studies^34^.

There are some limitations to the study. *First*, despite our inclusion of data from individuals of African-like ancestry, sample sizes of these discovery GWAS and the target AFR-like ABCD sample were small. Summary statistics for other global ancestral groups was absent. *Second*, data from Million Veterans Program, which over-represents older male veterans, are a major component of the SUD GWAS from which PRS were derived, potentially influencing associations; however, these GWAS appear to genetically correlate with GWAS from other samples (e.g., rg=0.66-1.00^35,36^), and our analyses of sex differences did not yield significant results. *Third*, there was little endorsement of substance use beyond alcohol sipping^37^, so we were unable to examine associations with substance use phenotypes beyond alcohol sipping and caffeine consumption. It is also possible that early substance experimentation is not strongly related to SUD genetic liability, and/or that it is better explained by genetic factors related to externalizing and sensation seeking^38,39^. *Fourth*, we elected to use Bonferroni correction, which given substantial inter-phenotype correlations, may be overly conservative. Because of this potential for false negatives, FDR-corrected and uncorrected p-values are also presented. *Fifth,* it is worth noting that interrogation of specific mechanisms that link SUD_PRS_ to individual behaviors (e.g., SUD_PRS_ influencing peer rule-breaking via gene-environment correlation) were beyond the scope of the current study, and alternative mechanisms to those proposed herein are also likely.

## Conclusions

By identifying associations between genetic liability to SUDs and conditions observable well before the onset of problematic (or any) substance use, the current PheWAS provides insights into childhood behaviors and environments that co-aggregate with and may be harbingers of future problems. While the current analyses do not test causal hypotheses regarding the role of these childhood phenotypes on later SUD, nor can PRS alone provide complete prognostic insight into multifactorial phenotypes like SUD, many of the associated phenotypes, such as caffeine consumption, screen time and social media use, sleep problems, and cigarette availability represent modifiable targets, interventions for which would broadly benefit youth health, and likely even more so in genetically susceptible youth.

## Acknowledgments

SEP, AA, and RB conceptualized the study. SEP, AJG, NRK, and APM curated and verified the data for analysis. SEP, DAAB, and ASH developed the R code. SEP conducted analyses and generated tables and figures. SEP, AA, and RB drafted the manuscript. SEP, AJG, NRK, APM, DAAB, ECJ, KHL, GD, DMB, ASH, AA, and RB critically reviewed the manuscript and take responsibility for the decision to submit the manuscript for publication.

## Code availability

Code is available upon request from the corresponding author.

## Data Availability

Data used in the preparation of this Article were obtained from the Adolescent Brain Cognitive Development (ABCD) study (https://abcdstudy.org), held in the NIMH Data Archive (NDA). This is a multisite, longitudinal study designed to recruit more than 10,000 children aged 9–10 years and follow them over 10 years into early adulthood. The ABCD data repository grows and changes over time.

## Inclusion & Ethics

Due to a dearth of GWAS in populations other than those genetically most similar to European and African reference populations, the present analyses were restricted to these individuals. The results may not generalize to other populations. In addition, the GWAS in African-like ancestry are smaller and thus less well-powered, likely limiting the discovery of phenotypes associated with SUD genetic risk in the African-like ancestry subsample.

The ABCD Study is a large sample of youth, which is an understudied population.

---

**Extended Data Figure 1.**
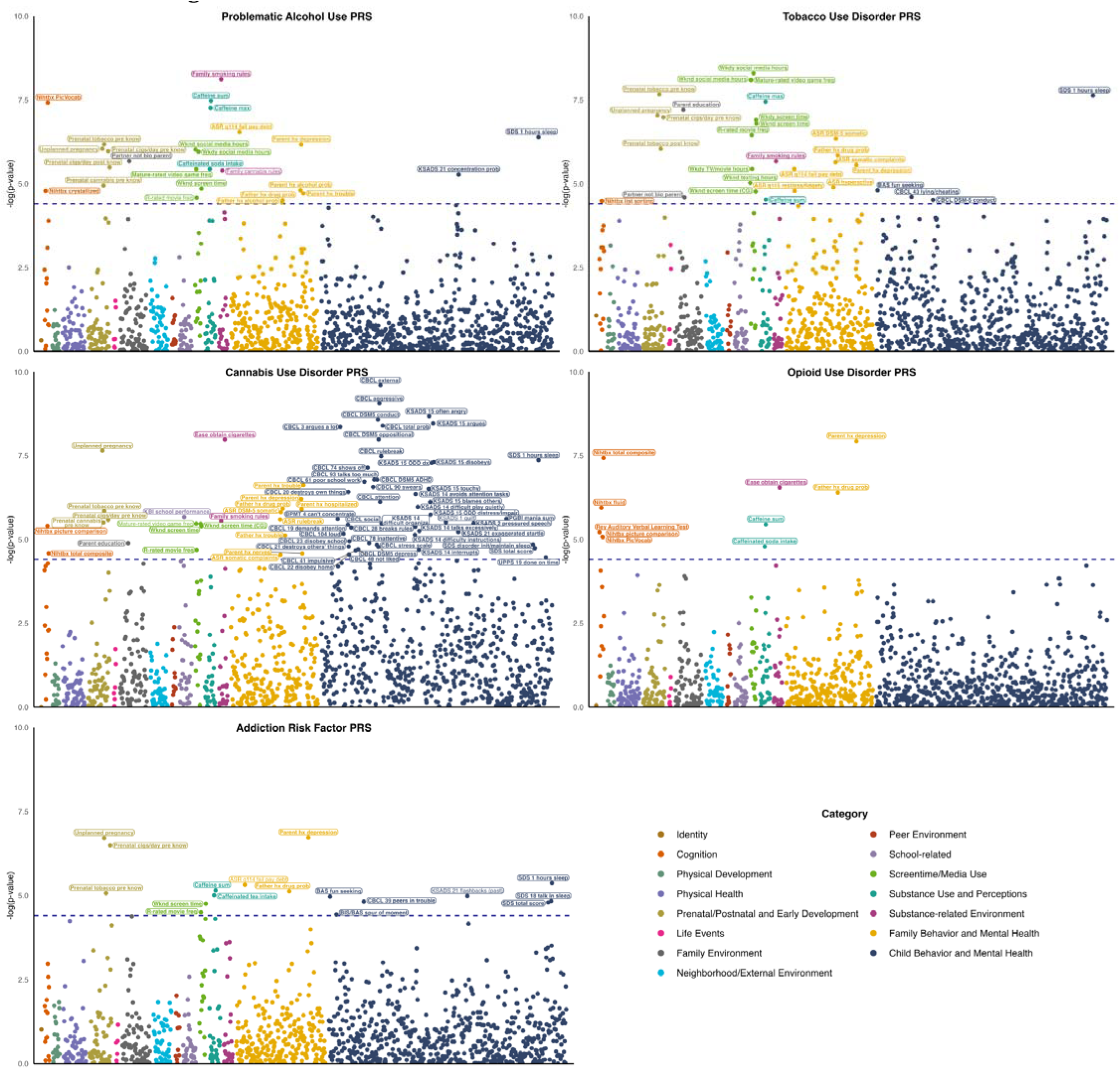
Note. Manhattan plots depicting primary PheWAS associations at the baseline wave among individuals genetically similar to European reference populations. The 1,271 examined phenotypes are spread along the x-axis, and the y-axes represent the -log10 of the uncorrected p values of associations between each PRS and each phenotype. The points are color-coded by phenotype domain. The dashed lines indicate the Bonferroni-significant threshold (i.e., -log10(0.05/1271) = 4.405). Full results are located in Supplemental Table 9.

**Extended Data Figure 2.**
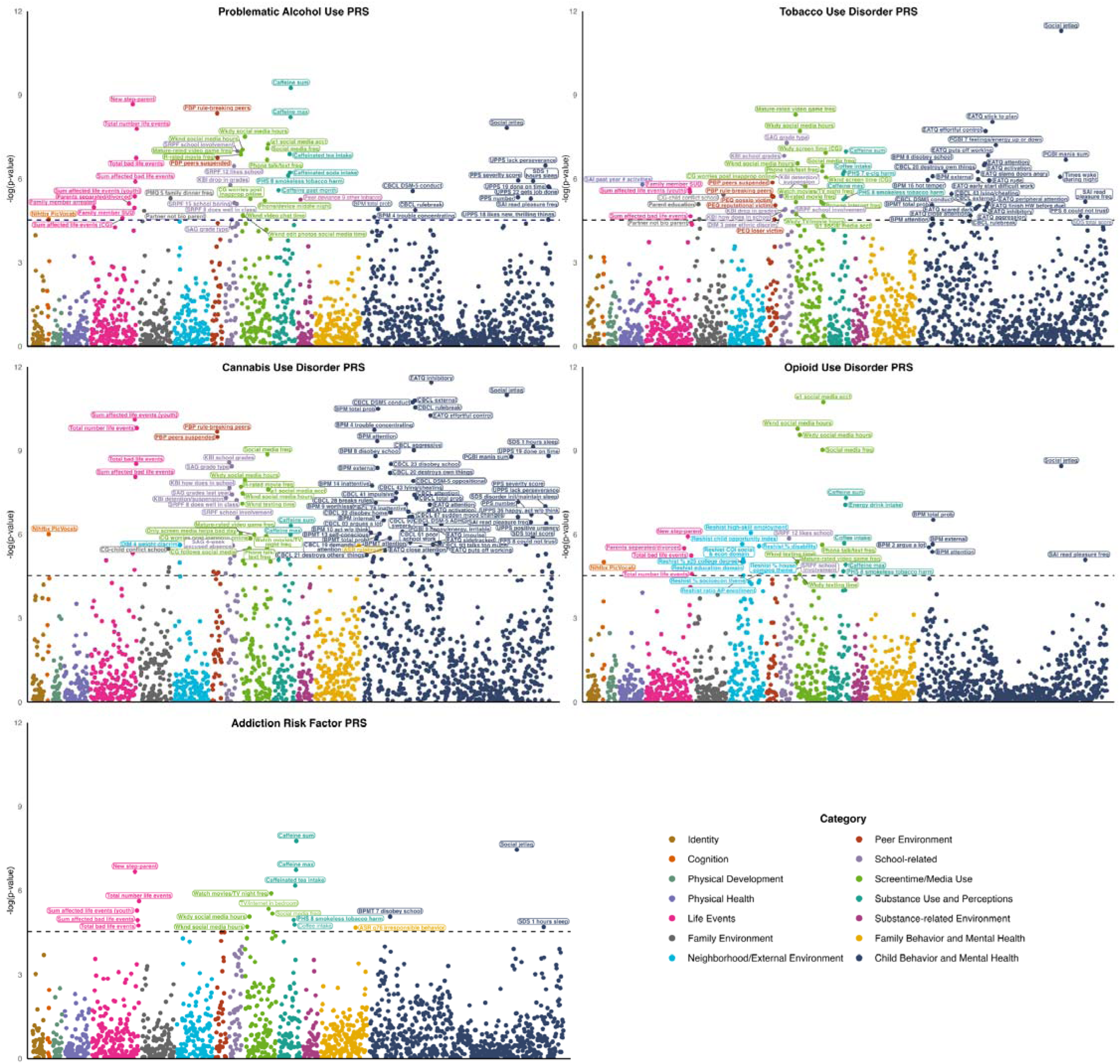
Note. Manhattan plots depicting primary PheWAS associations at the two-year follow-up wave among individuals genetically similar to European reference populations. The 1,697 examined phenotypes are spread along the x-axis, and the y-axes represent the -log10 of the uncorrected p values of associations between each PRS and each phenotype. The points are color-coded by phenotype domain. The dashed lines indicate the Bonferroni-significant threshold (i.e., -log10(0.05/1697) = 4.53). Full results are located in Supplemental Table 10.

**Extended Data Figure 3.**
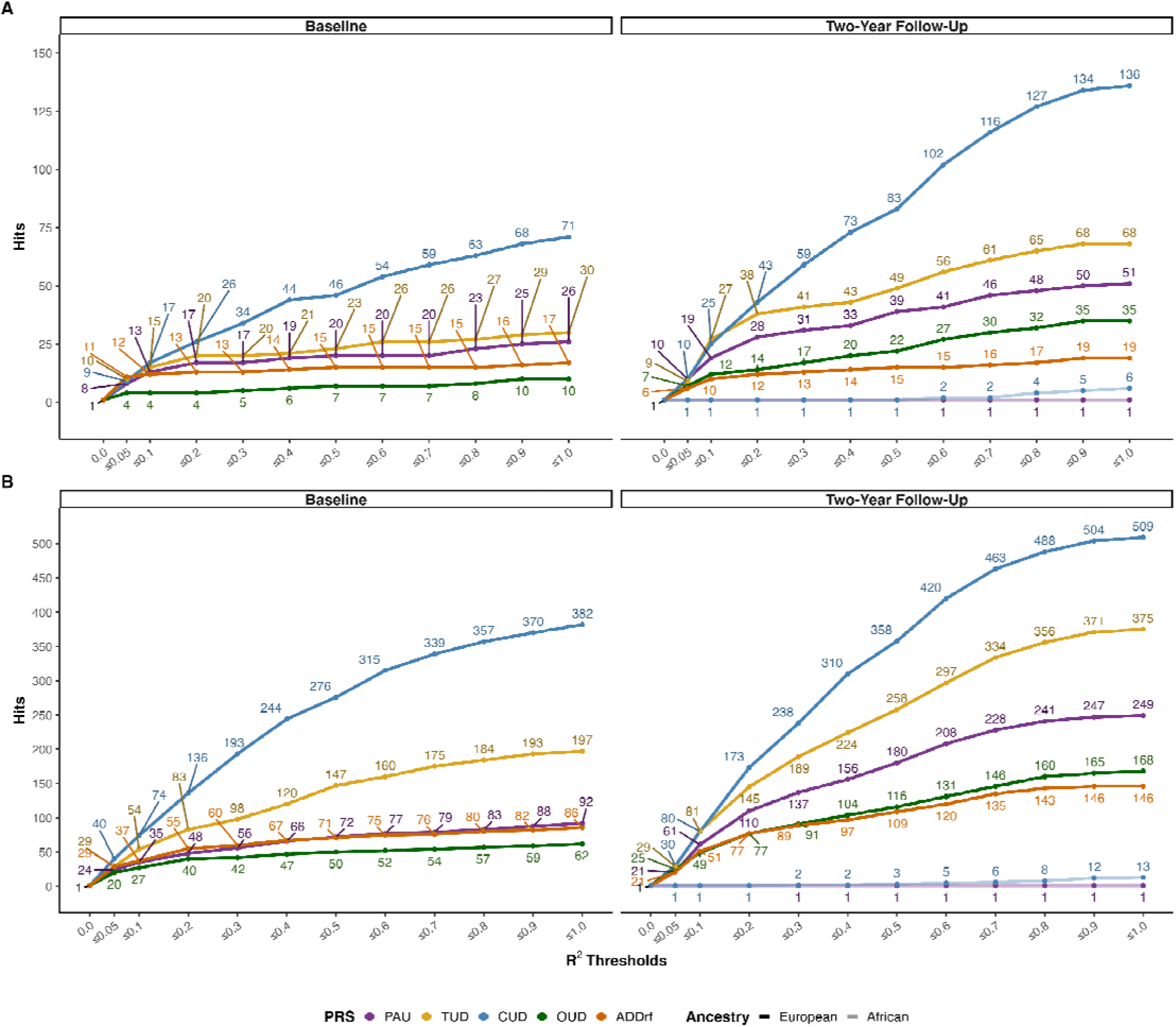
Pruning significant associations according to correlations among phenotypes and strength of association between each phenotype and each PRS. Pseudo R2 values were obtained for each pair of phenotypes significantly (A, Bonferroni-corrected; B, FDR-orrected) associated with each PRS. At each R2 threshold, the phenotype with the higher (i.e., less significant) p-value within each henotype pair whose R2 was above the threshold was removed. The following R2 thresholds were examined: 0.05, 0.1, 0.2, 0.3, 0.4, 0.5, 0.6, 0.7, 0.8, 0.9, 1.0, with 1.0 including all significantly associated phenotypes. Colors denote different PRS. More transparent lines denote associations in individuals most genetically similar to African reference populations; these are only included in the Two-Year Follow-Up plots given the lack of significant associations observed at Baseline. PAU = Problematic Alcohol Use; TUD = Tobacco Use Disorder; CUD = Cannabis Use Disorder; OUD = Opioid Use Disorder; ADDrf = Addiction Risk Factor; AUD = Alcohol Use Disorder.

**Extended Data Figure 4.**
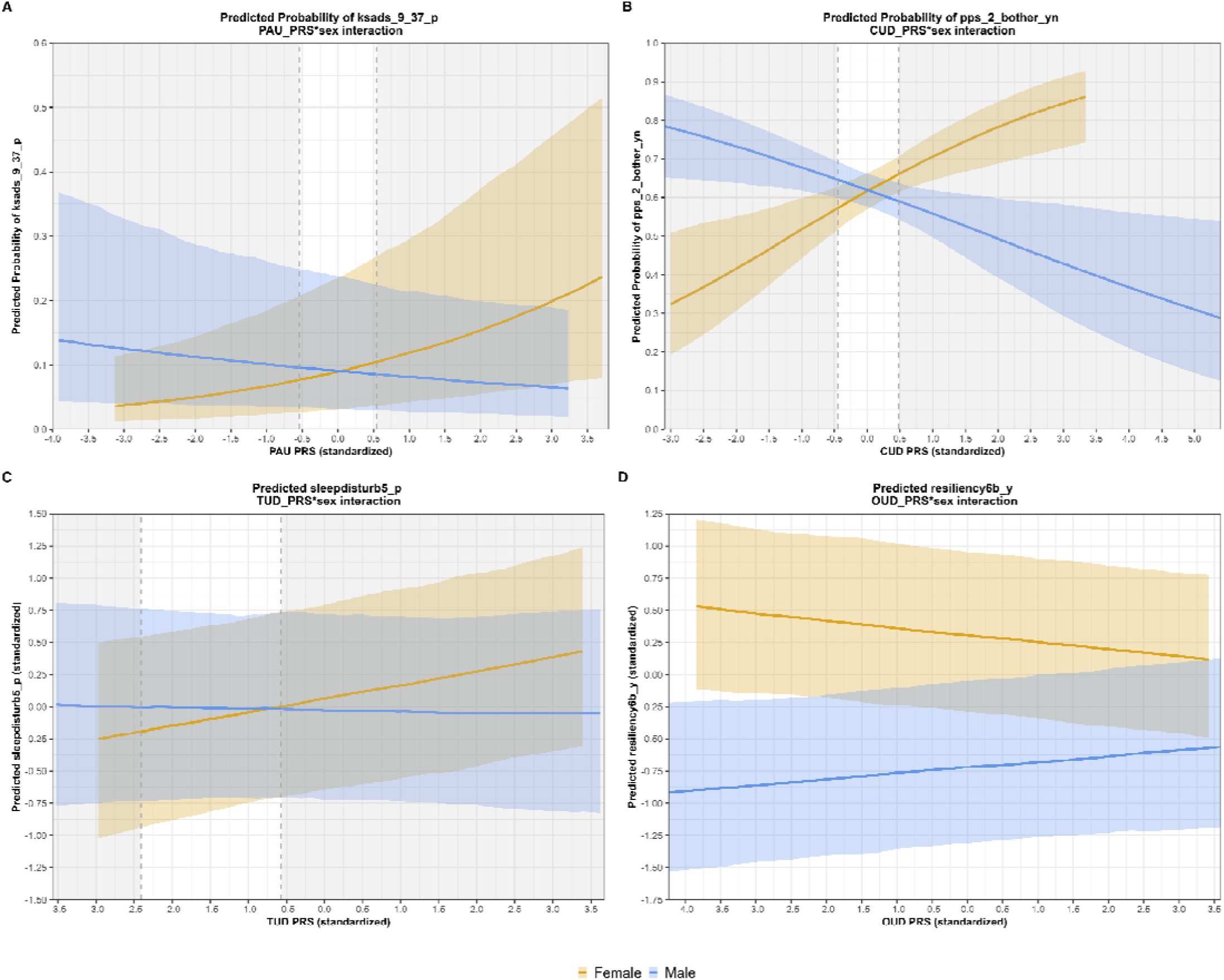
Note. Interactions between SUD PRS and biological sex in the sample of participants genetically similar to European reference populations. Confidence intervals around the predicted probabilities are shown in yellow (female) and blue (male). Gray shaded areas indicate Johnson-Neyman intervals for which the interaction is significant. A. Interaction between Problematic Alcohol Use (PAU) PRS and biological sex in association with ksads_9_37_p (past active avoidance of a phobic object). Confidence intervals around the predicted probabilities are shown in yellow (female) and blue (male). The simple slope for female participants is statistically significant (B=0.31, p=5.12e-05), and the simple slope for male participants is not (B=-0.12, p=0.09). The interaction is significant outside of the range -0.54 to 0.54. B. Interaction between Cannabis Use Disorder (CUD) PRS and biological sex in association with pps_2_bother_yn (bothered by hearing strange sounds). The simple slopes for female (B=0.42, p=2.16e-06) and male (B=-0.24, p=0.02) participants are both statistically significant but opposite in direction. The interaction is significant outside of the range of - 0.45 to 0.48. C. Interaction between Tobacco Use Disorder (TUD) PRS and biological sex in association with sleepdisturb5_p (feeling anxious or disturbed when falling asleep). The simple slope for female participants is statistically significant (B=0.10, p=5.63e-06), and the simple slope for male participants is not (B=-0.02, p=0.41). The interaction is significant outside of the range of -2.41 to -0.57. D. Interaction between Opioid Use Disorder (CUD) PRS and biological sex in association with resiliency6b_y (number of close friends that are girls). The simple slope for male participants is statistically significant (B=0.08, p=3.22e-07), and the simple slope for female participants is not (B=-0.02, p=0.26). The Johnson-Neyman Intervals for which the interaction is significant fall outside of the range of observed data and thus are not shown here.

**Extended Data Figure 5.**
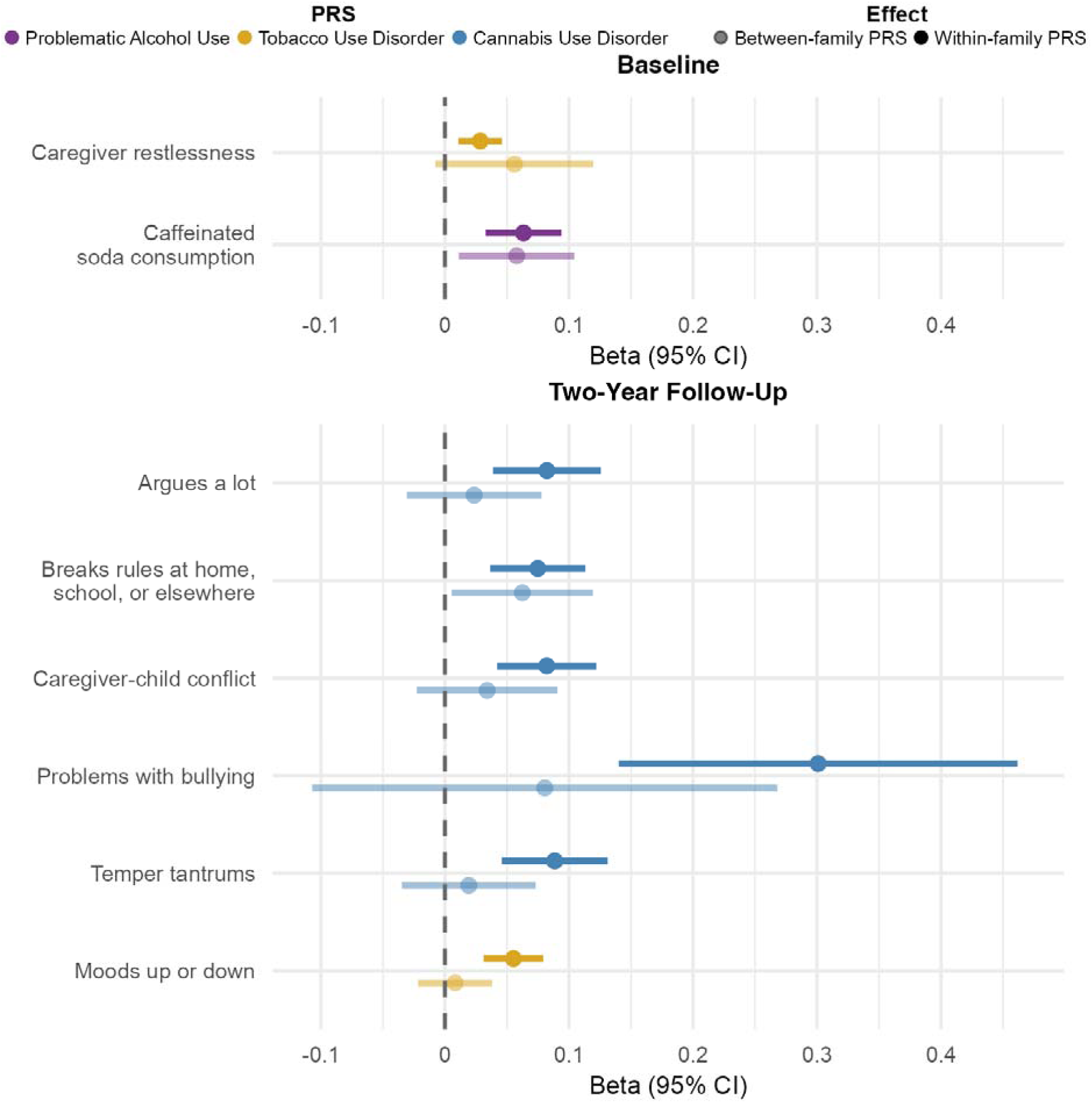
*Note.* Within-family analyses in European-like genetic ancestry. PRS were decomposed into between- and within-family variance and entered into mixed effects models simultaneously as predictors of phenotypes that were significantly associated with the PRS following Bonferroni correction. Depicted here are the Bonferroni-corrected significant within-family PRS associations (in the darker color), alongside the respective between-family PRS effect (lighter color). Point estimates reflect standardized betas, and error bars represent 95% confidence intervals.

**Extended Data Figure 6.**
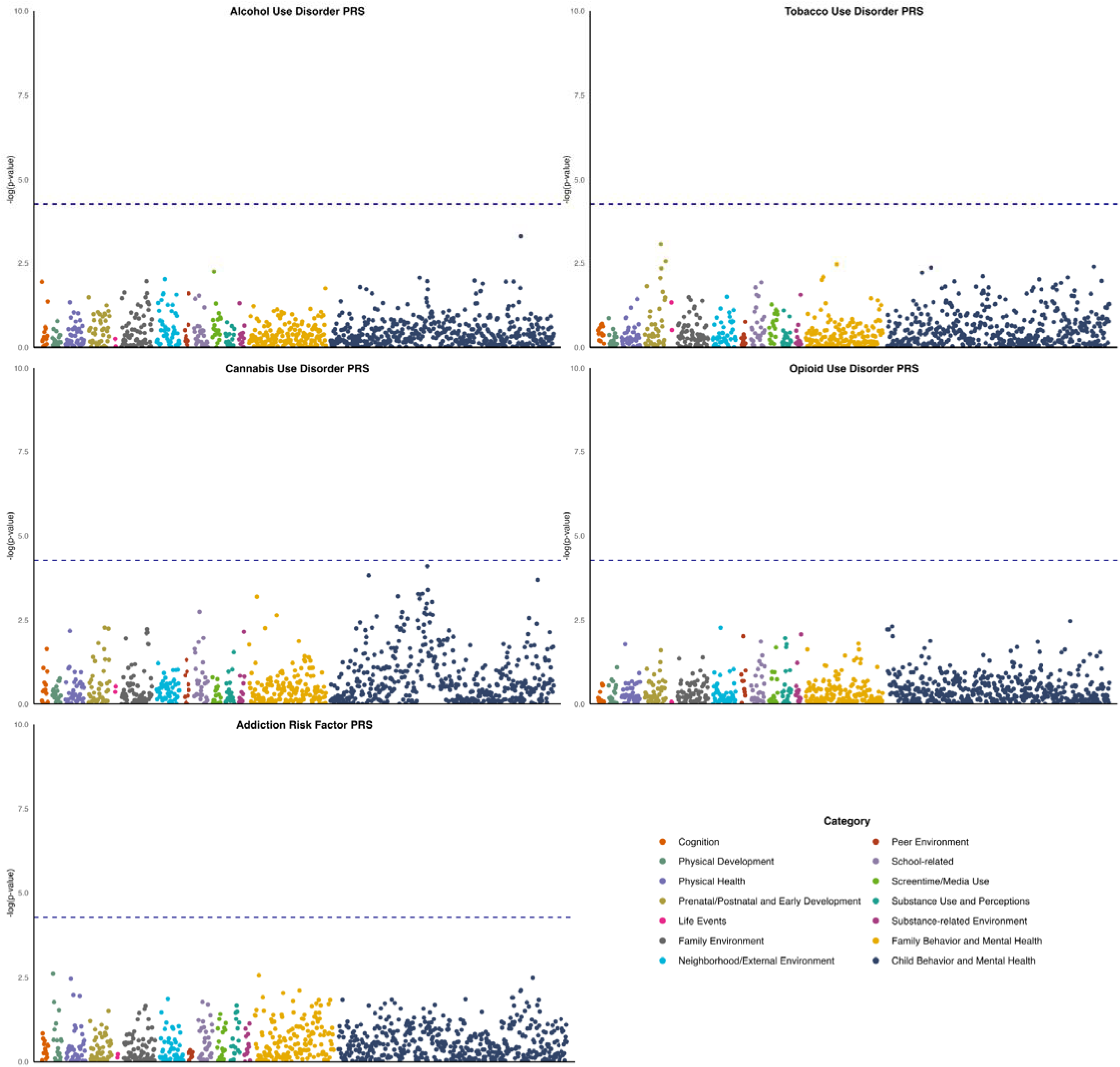
*Note*. Manhattan plots depicting primary PheWAS associations at the baseline wave among individuals genetically similar to African reference populations. The 940 examined phenotypes are spread along the x-axis, and the y-axes represent the -log_10_ of the uncorrected *p* values of associations between each PRS and each phenotype. The points are color-coded by phenotype domain. The dashed lines indicate the Bonferroni-significant threshold (i.e., -log_10_(0.05/940) = 4.27). Full results are located in **Supplemental Table 21**.

**Extended Data Figure 7.**
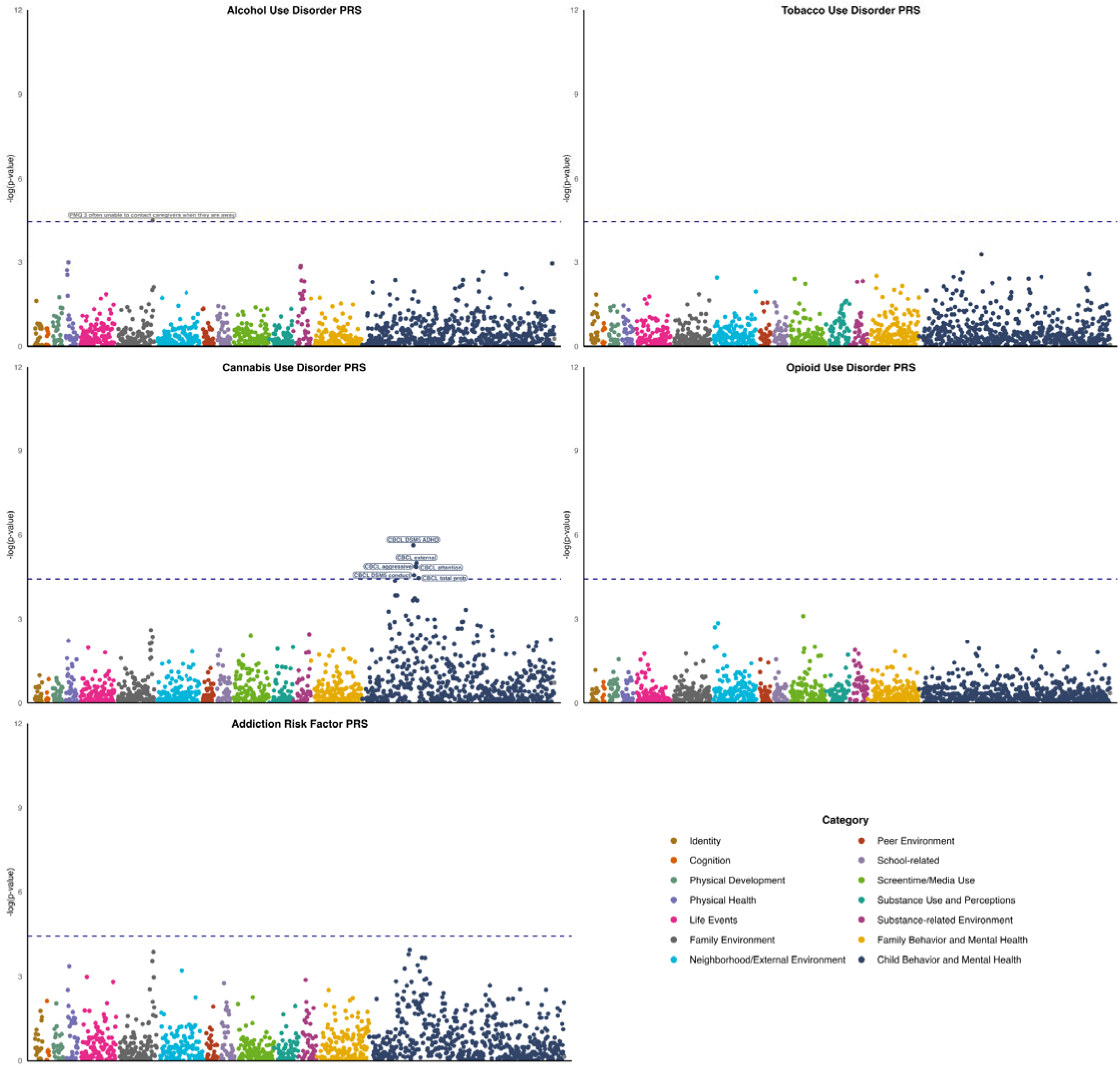
*Note*. Manhattan plots depicting primary PheWAS associations at the two-year follow-up wave among individuals genetically similar to African reference populations. The 1,357 examined phenotypes are spread along the x-axis, and the y-axes represent the -log_10_ of the uncorrected *p* values of associations between each PRS and each phenotype. The points are color-coded by phenotype domain. The dashed lines indicate the Bonferroni-significant threshold (i.e., -log_10_(0.05/1,357) = 4.43). Full results are located in **Supplemental Table 22**.

**Extended Data Figure 8.**
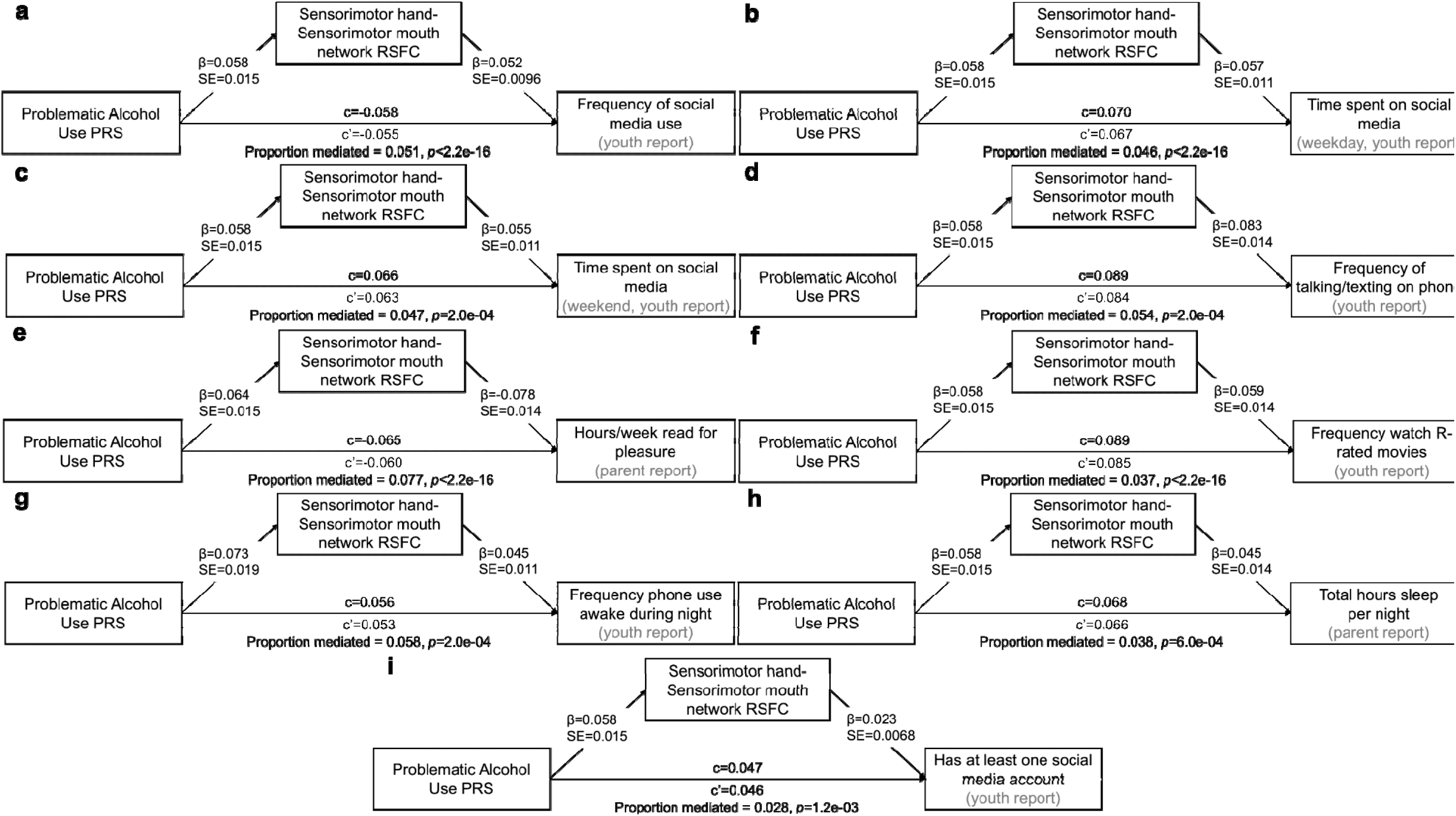
Indirect effects of Problematic Alcohol Use PRS on screen time, reading, and sleep phenotypes through resting state functional connectivity (RSFC) between the sensorimotor hand and sensorimotor mouth networks. c = total effect; c’ = direct effect. All indirect effects shown are statistically significant following Bonferroni correction. Results are also shown in **Supplemental Table 35.** These results are specific to individuals genetically similar to European reference populations.

**Extended Data Figure 9.**
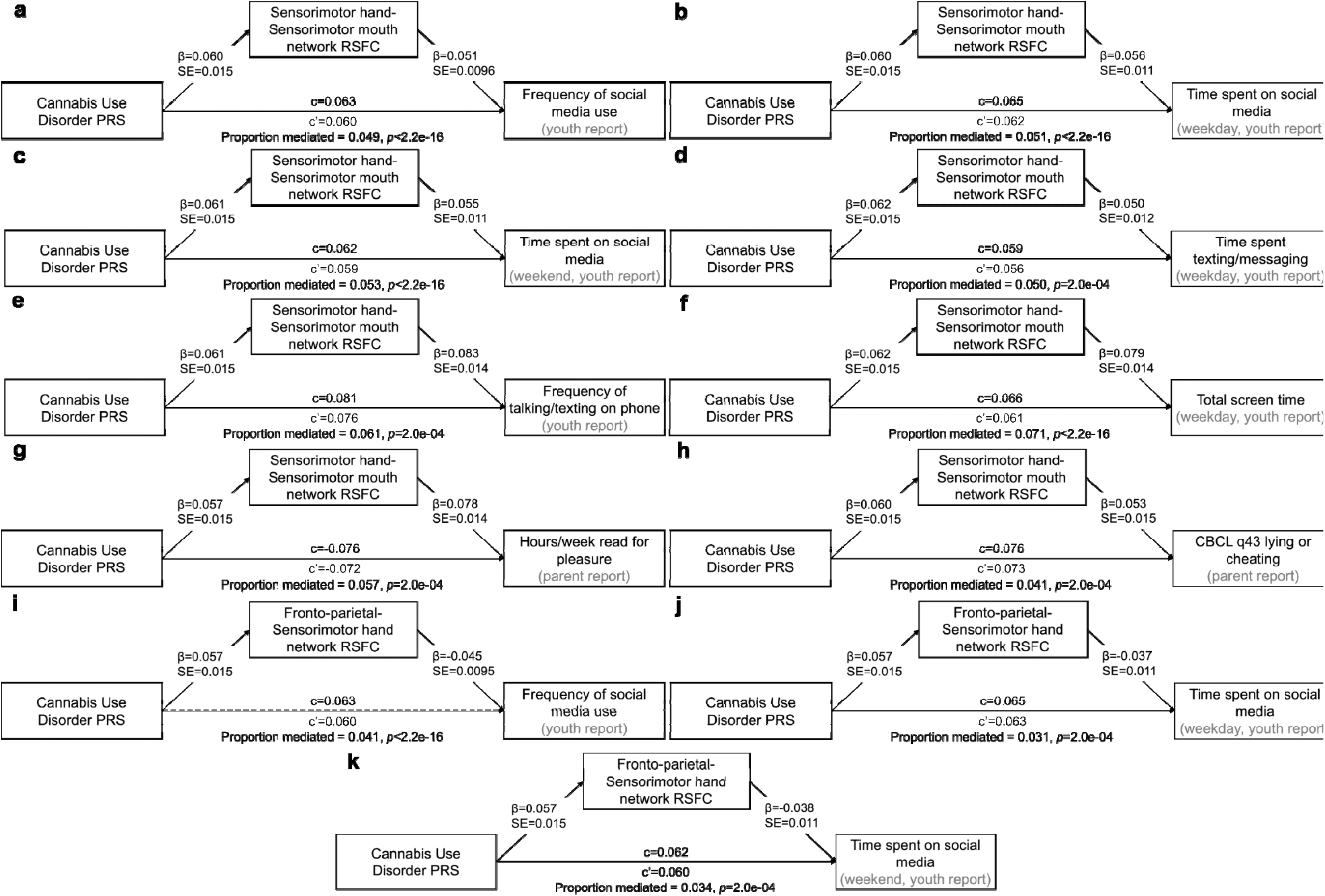
Indirect effects of Cannabis Use Disorder PRS on screen time, reading, and lying/cheating phenotypes through resting state functional connectivity (RSFC) between the sensorimotor hand and sensorimotor mouth networks and between the fronto-parietal and sensorimotor hand networks. c = total effect; c’ = direct effect. All indirect effects shown are statistically significant following Bonferroni correction. Results are also shown in **Supplemental Table 35.** These results are specific to individuals genetically similar to European reference populations.

**Extended Data Figure 10.**
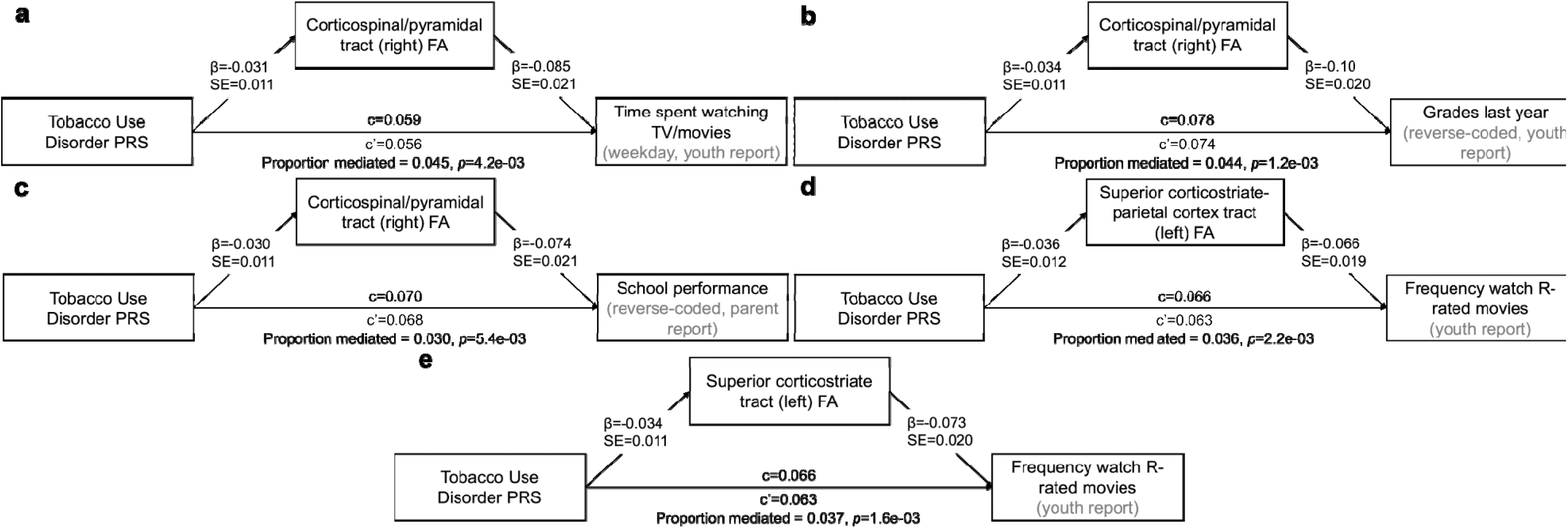
Indirect effects of Tobacco Use Disorder PRS on screen time and school performance phenotypes through fractional anisotropy in the corticospinal/pyramidal, superior corticostriate-parietal cortex, and superior corticostriate white matter tracts. c = total effect; c’ = direct effect. All indirect effects shown are statistically significant following Bonferroni correction. Results are also shown in **Supplemental Table 35.** These results are specific to individuals genetically similar to European reference populations.

## Footnotes

1 ORs <1 were converted to be ≥1 to facilitate interpretable range descriptions in text.

