## Supplementary material for "Youth Correlates of Genetic Liability to Substance Use Disorders": Methods

**Participants**	**2**

**Non-imaging phenotypes	2**

**Imaging phenotypes	2-3**

**Polygenic scores	3-4**

**Genotyping, quality control, and imputation	3-4**

**Polygenic score computation	4**

**Non-imaging analyses	4-5**

**Imaging analyses	5-6**

**Secondary analyses	6-9**

**Substance-naive PheWAS	6**

**Simultaneous SUD PRS PheWAS	6**

**Phenotypic pruning	6-7**

**PRS-by-time moderation	7**

**Sex moderation	7**

**Within-family PRS effects	7**

**PheWAS adjusted for phenotypic externalizing behavior	8**

**Indirect effects through neuroimaging-derived phenotypes	8**

**Cross-ancestry meta-analysis	8-9**

**References	10-12**

**Participants.** The Adolescent Brain Cognitive Development^SM^ (ABCD) Study is a longitudinal study of adolescent development in the United States^1^[. Our analyses were restricted to individuals whose genomic ancestry resembled African](https://abcdstudy.org/sites/abcd-sites.html) (n=1,584; mean±SD age = 9.90±0.61 years, 50.0% female) [and European (](https://abcdstudy.org/sites/abcd-sites.html)n=5,556, 9.93 ±0.63 years, 47.0% female) [reference populations (hereafter referred to as African- [AFR] and European-like [EUR] genetic ancestry, respectively; **Table 1**). Due to the lack of well-powered GWASs in other ancestral populations and](https://abcdstudy.org/sites/abcd-sites.html) challenges with cross-ancestry portability of summary data^2,3^, we did not analyze data from other genetic ancestries (n=3,959). A trans-ancestral meta-analysis is presented in the **Supplement**. Parental/caregiver written and child verbal informed consent/assent were given for a protocol approved by a centralized institutional review board and those at each data collection site (n=21; <https://abcdstudy.org/sites/abcd-sites.html>).

***Non-imaging phenotypes.*** A total of 940 and 1,271 baseline phenotypes and 1,357 and 1,697 follow-up phenotypes were included for AFR and EUR individuals, respectively. The assessments/measures from which these items were derived as well as any re-coding notes are listed in **Supplemental Tables 1-8** (including tables specific to substance-naive subsamples). Briefly, 6,194 and 10,156 non-imaging phenotypes were considered at baseline and follow-up, respectively. All phenotypes were first examined for relevance, and irrelevant items (e.g., redundant items, such as raw and t-scores; administrative items, such as assessment device and items indexing the number of questions answered or missing, language of interview) were removed (1,571 at baseline, 5,747 at follow-up). Then, following previous studies ^4,5^, variables with fewer than 100 data points or fewer than 100 endorsements of minority categories of categorical variables were removed, separately for each ancestry group, leaving 940 and 1,271 baseline phenotypes and 1,357 and 1,697 follow-up phenotypes--including dummy-coded categories of categorical variables--included for AFR and EUR individuals, respectively. These data curation procedures were conducted by multiple clinically trained investigators (SEP, NRK, AJG) to ensure replicability and provide consensus about relevance and recoding.

Phenotypes were grouped into domains for descriptive and interpretative purposes: **1)** Identity (n_phenotypes_=0-40), **2)** Cognition (n_phenotypes_=8-13), **3)** Physical Development (n_phenotypes_=18-31), **4)** Physical Health (n_phenotypes_=39-84), **5)** Prenatal/Postnatal and Early Development (n_phenotypes_=40-60 [baseline only]), **6)** Life Events (n_phenotypes_=2-162), **7)** Family Environment (n_phenotypes_=64-111), **8)** Neighborhood/External Environment (n_phenotypes_=48-123), **9)** Peer Environment (n_phenotypes_=10-38), **10)** School-related (n_phenotypes_=29-45), **11)** Screentime/Media Use (n_phenotypes_=18-106), **12)** Substance Use and Perceptions (n_phenotypes_=19-75), **13)** Substance-related Environment (n_phenotypes_=12-52), **14)** Family Behavior and Mental Health (n_phenotypes_=137-239), and **15)** Child Behavior and Mental Health (n_phenotypes_=464-663; **Supplemental Tables 1-8**).

***Imaging phenotypes.*** Imaging phenotypes included cortical and subcortical volume, thickness, and surface area; resting-state functional connectivity; and white matter fractional anisotropy and mean diffusivity. A detailed description of the imaging acquisition procedures and processing and analysis methodology in ABCD can be found elsewhere ^6,7^.

Structural MRI (sMRI) metrics were selected from the Desikan atlas: 15 global metrics (total and bilateral cortical volume and surface area, mean cortical thickness, total subcortical gray matter volume, intracranial and whole brain volume, and supratentorial volume, as well as bilateral cerebral white matter volume. An additional 239 regional sMRI metrics (i.e., 34 cortical regions*2 hemispheres*3 metrics (i.e., volume, surface area, thickness) + 35 subcortical regions (i.e., accumbens, thalamus, caudate, putamen, pallidum, hippocampus, amygdala, ventral diencephalon, brainstem, cerebellum, ventricles, cerebrospinal fluid, corpus callosum segments) were extracted for potential follow-up. Only sMRI data that passed QC tests (Freesurfer QC criteria and had at least one T1 scan that passed sMRI cortical surface reconstruction QC criteria; EUR n=5,517; AFR n=1,543) were retained ^7^.

For diffusion MRI (dMRI), 2 global metrics (total fractional anisotropy and mean diffusivity for all fibers) and 37 regional metrics each for FA and MD were included. Only dMRI data that passed QC tests (passed Freesurfer QC criteria, had at least one dMRI scan that passed QC criteria, and did not have an overall dMRI QC score of 0 based on B0 warp, registration, image quality, and segmentation; EUR n=5,294; AFR n=1,448) were retained ^7^.

Resting state functional connectivity (rs-fMRI) metrics were acquired from four 5-minute resting-state BOLD scans, with pair-wise correlations computed for ROIs within functionally-defined parcellations (i.e., Gordon networks). The Fisher Z-transform of the correlation values were examined within each of the 13 Gordon networks and across 78 additional between-network rs-fMRI metrics). Only rs-fMRI data that passed QC tests (passed Freesurfer QC criteria and had at least one rs-fMRI scan that passed QC criteria; EUR n=5,353; AFR n=1493) were retained ^7^.

**Polygenic Scores.** PRS-CS (European ancestry^8^), PRS-CSx (African ancestry^9^), and PLINK v2.0^10^ were used to compute polygenic risk scores (PRS) using the largest available published GWASs (**Table 2**, **Supplement**) of: **1)** Problematic alcohol use (PAU)/Alcohol Use Disorder (AUD^11^), 2) **2)** Tobacco Use Disorder (TUD^12^), **3)** Cannabis Use Disorder (CUD^13^), **4)** Opioid Use Disorder (OUD^14^), and **5)** Addiction Risk Factor (ADDrf^4^; **Table 2)**.

**Polygenic Risk Scores**

***Genotyping, Quality Control, and Imputation.*** Saliva samples were genotyped on the Smokescreen array ^15^ by the Rutgers University Cell and DNA Repository (now incorporated with other companies as Sampled; https://sampled.com/). The Rapid Imputation and COmputational PIpeLIne for Genome-Wide Association Studies (RICOPILI^16^) was used to perform quality control (QC) on the 11,099 individuals with available ABCD Study phase 3.0 genotypic data, using RICOPILI’s default parameters (genotyping call rate >98%, inbreeding coefficient (F) < ±0.2, sex checks). The 10,585 individuals who passed these QC checks were matched to broad self-reported racial groups using the ABCD Study parent survey. Of the 6,787 parents/caregivers indicating that their child’s race was only “white,” 5,561 of those individuals did not endorse any Hispanic ethnicity/origin. Of the 1,675 parents/caregivers who indicated that their child’s race was only “Black,” 1,584 did not endorse any Hispanic ethnicity/origin.

Next, variants were filtered to exclude those with missingness >2% and HWE p-value <1e-06. Using data from unrelated individuals (pi-hat ≤ 0.2) and an LD pruned set of common (MAF>0.05) and non-palindromic SNPs (and excluding MHC and chromosome eight inversion region), principal components analysis (PCA) was performed in RICOPILI using EIGENSTRAT^17^ to confirm the genetic ancestry of these individuals. This was done by merging the ABCD Study data with the 1000 Genomes reference panel, computing the mean and standard deviations for the 1000 Genomes ancestry populations for the top 6 PCs, and establishing that the previously identified group participants were genetically similar to the 1000 Genomes European and African reference panels by assessing whether they fell within 3SDs of the mean for the top 6 PCs within the respective 1000 Genomes reference population. After another round of QC on these subsamples, 5,556 individuals of European ancestry and 1,584 individuals of African ancestry were retained in the analyses. Each ancestry subset was then imputed to the TOPMed imputation reference panel^17^. Imputation dosages were converted to best-guess hard-called genotypes, and only SNPs with Rsq > 0.8, MAF > 0.01, missingness < 0.1, and HWE p-values >1E-06 were kept for PRS analyses.

***Polygenic Scoring.*** PRS-CS^17^, PRS-CSx^9^, and PLINK v2.0^10^ were used to compute polygenic risk scores (PRS) for the substance use disorder phenotypes described below. PRS-CS is a Bayesian polygenic prediction method that applies a continuous shrinkage prior to SNP effect estimates and infers posterior SNP weights. PRS-CSx is an extension of PRS-CSx that integrates summary statistics and LD reference panels from multiple ancestral groups to enable more accurate polygenic prediction in populations for which sufficiently powered GWAS may not exist. Parameters for both PRS-CS and PRS-CSx included default phi parameters and n_iter = 10,000, n_burnin = 5,000; PRS-CS used the auto function, and in PRS-CSx, the meta flag was set to TRUE. The PLINK –score command was used to calculate PRS for each individual by summing all variants weighted by the inferred posterior effect size.

The following SUD GWAS summary statistics were used: Problematic Alcohol Use (PAU; EUR N=903,147^11^), Alcohol Use Disorder (AUD; AFR N=122,571, supplemented with N=753,248 EUR^11^), Tobacco Use Disorder (TUD; AFR N=114,420; EUR N=739,895^12^), Cannabis Use Disorder (CUD; AFR N=123,208; EUR N=886,025^13^), and Opioid Use Disorder (OUD; AFR N=84,877; EUR N=554,186^14^), and Addiction Risk Factor (ADDrf; AFR N=92,630; EUR N=1,025,550^4^; **Table 2**).

**Non-Imaging Analyses**

Primary analyses examined associations between each PRS and all curated phenotypes, separately for each ancestry group and wave of data collection. Variables were examined for skew, and all continuous variables with skew ≥|1.96| were winsorized to 3SDs. Variables with skew still ≥|1.96| were subjected to inverse normal transformation. All other continuous variables were standardized (i.e., Z-scored) prior to analyses to enable consistent interpretation of effect sizes. Linear (for continuous outcomes) and generalized linear (for dichotomous outcomes) mixed effects models were fit to the data using the lme4 R package^18^, with random intercepts for family and data collection site and the following fixed effects covariates:

**1-10) First 10 ancestral principal components (PCs).** As described above, ancestral PCs were computed separately for those genetically similar to European (EUR) and African (AFR) reference populations. The first 10 of these were standardized and included as fixed effects covariates in all models that included PRS.

**11) Age.** Age (in months) was standardized separately at each assessment wave.

**12) Sex.** Biological sex at birth was caregiver-reported and had two levels: male and female.

**13) Visit type.** For analyses using data from the two-year follow-up, part of which coincided with the COVID-19 pandemic, visit type was included to account for any differences across in-person, hybrid, or remote assessments.

***Statistical significance.*** Statistical significance was determined via Bonferroni correction for the number of tests (i.e., 940 and 1,271 tests at baseline, 1,361 and 1,697 tests at follow-up), separately for each PRS. False discovery rate (FDR)-corrected p-values are also presented, as the Bonferroni approach may be overly stringent given correlations among phenotypes. Models that did not converge due to insufficient variability in covariates or random effects were modified to exclude problematic parameters (e.g., sex was removed from pubertal development models where phenotypes were assessed separately for boys and girls).

**Imaging Analyses**

Linear mixed-effects models were also used to test associations between each of the PRS and imaging metrics, which were also standardized and scaled. In addition to the covariates noted above, the following were included:

**MRI scanner type.** A variable denoting whether the MRI was Siemens, Philips, or GE was included.

**Motion.** For analyses including resting-state functional connectivity (RSFC) or diffusion-weighted MRI (DWI) metrics, the respective motion correction covariate was included.

**Global metrics.** For analyses examining regional cortical volume, surface area, and thickness metrics, the respective global metric was added as a covariate (i.e., total cortical volume, total cortical surface area, mean cortical thickness). Total subcortical gray matter volume was included as a covariate in analyses examining regional subcortical area metrics. Mean fractional anisotropy and mean diffusivity across all fiber tracts were added as covariates to analyses examining fractional anisotropy and mean diffusivity across individual fiber tracks, respectively.

***Statistical significance.*** Bonferroni and FDR correction were applied to 1) all p-values (n=421) and 2) separately to all p-values within each modality (i.e., global, cortical volume, surface area, thickness, subcortical volume, rs-fMRI, dMRI FA, dMRI MD) within each PRS model.

Imaging analyses were also repeated in the substance-naive subsamples.

**Secondary analyses**

**1) Substance-naive PheWAS**. The data were restricted to individuals who had not yet initiated any substance use (i.e., “substance-naive”). Phenotypes significantly associated with SUD PRS in this sample may reflect developmental expression of SUD risk prior to onset of use that may mechanistically link genetic liability to future initiation and/or substance use-related problems. They could also reflect protective factors against early initiation or future use and related problems. These hypotheses will be crucial to test as this sample ages into substance initiation and regular use.

Substance use initiation was defined as endorsement of any substance use (e.g., alcohol “sipping,” nicotine or cannabis “puffing,” use of any substance through any form or route of administration) outside of the context of religious ceremonies. Substance-naive individuals had non-missing substance use data at the point of assessment (i.e., baseline, follow-up) and did not report any substance use (including during religious ceremonies) at any point up to and including the current assessment. For more information, see^19^. There were 1,411 and 4,289 substance-naive individuals of African and European ancestry, respectively, at baseline, and 1,125 and 3,704 substance-naive individuals at follow-up. The substance-naive samples did not differ from the full samples by sex, age, household income, or caregiver sex (*p*≥0.079).

Primary PheWAS analyses (i.e., primary PheWAS and imaging PheWAS in both waves and genetic ancestry groups) were repeated in these substance-naive samples. P-value correction (Bonferroni and FDR) was used to adjust for the number of tests (the number of Bonferroni-significant phenotypes from primary analyses that also passed quality control criteria in the substance-naive subsamples).

**2) Simultaneous SUD PRS PheWAS.** The four non-ADDrf SUD PRS were entered into PheWAS analyses simultaneously to identify potentially independent SUD PRS associations. ADDrf PRS was excluded to avoid potential over-correction for shared variance, as the Addiction Risk Factor was modeled to capture overlapping genetic variance across the four SUDs included here. Bonferroni and FDR correction were used to adjust p-values across all phenotypes, separately by each SUD PRS and by wave.

**3) Phenotypic pruning.** Given interrelatedness among phenotypes, PheWAS-significant phenotypes were pruned based on their degree of overlap as determined by pseudo R^2^ (Paul et al., 2024). First, linear mixed-effects models were used to estimate bivariate pseudo R^2^ values (lme4^18^ and MuMIn^20^ R packages) between each PheWAS-significant phenotype, with random intercepts for family and research site. Second, for each R^2^ threshold examined (0.05, 0.1, 0.2, 0.3, 0.4, 0.5, 0.6, 0.7, 0.8, 0.9, 1.0), the phenotype with the higher (i.e., less significant) PheWAS p-value within each phenotype pair whose R^2^ value was at or above the R^2^ threshold was iteratively removed until no remaining R^2^ values met that threshold.

**4) PRS-by-time moderation.** For each of the 802 and 619 phenotypes assessed at both baseline and follow-up for European and African ancestry, respectively, each PRS (separately) was interacted with time point (baseline and two-year follow-up), controlling for sex, person-mean age, the first 10 ancestral PCs, and all predictor-by-covariate interactions, and including random intercepts for individual, family, and site. The PRS-by-time estimate can be interpreted as the extent to which PRS is associated with phenotypic change from baseline to follow-up. Bonferroni correction was applied to the 802and 619 interaction p-values, separately for each PRS.

**5) Sex moderation.** To test whether biological sex at birth moderated associations between each PRS and phenotype, PRS-by-sex interactions were examined for all phenotypes with sufficient data from both boys and girls (i.e., sex-specific pubertal development variables were excluded, as were any dichotomous variables for which there were not at least 100 endorsements of each level from both boys and girls, as well as any continuous phenotypes for which there were not at least 100 non-missing data points for both boys and girls). All predictor-by-covariate interactions (e.g., PRS-by-age, sex-by-age) were included to appropriately control for confounding^21^). The PRS-by-sex interaction p-values were subject to Bonferroni correction for the number of tests within each PRS (i.e., 1,017 and 705 phenotypes at baseline, 1,369 and 1,109 phenotypes at follow-up for individuals of European and African ancestry, respectively). Simple slopes and Johnson-Neyman intervals (using the sim_slopes() and johnson_neyman() functions, respectively, from the “interaction” package^22^) were computed for all statistically significant interactions.

**6) Within-family PRS effects.** PRS associations reflect a combination of direct (i.e., causal) genetic effects and confounding due to population stratification, assortative mating, and/or passive gene-environment correlation. Within-family analyses, wherein the PRS is decomposed into between- (family-mean PRS) and within-family (each individual’s deviation from the family-mean PRS) variance, offer a way to exploit family-based designs to more closely approximate whether PRS associations may plausibly reflect causal genetic effects on outcomes ^23–25^. For these analyses, the sample was restricted to families with at least 2 siblings participating in the ABCD study (baseline EUR n=1809 [n=893 families], follow-up EUR n=1655 [n=827 families]; follow-up AFR n=316 [n=156 families]; baseline AFR excluded due to lack of significant baseline associations). Both family-mean PRS and each sibling’s deviation from that family-mean PRS were entered into mixed effects models to estimate the between- and within-family PRS effects, with the latter more closely reflecting any direct genetic effects. Only phenotypes that were previously identified as significantly associated with each PRS, following Bonferroni correction, and that passed quality control criteria within the smaller sibling subsamples (i.e., at least 100 data points or at least 100 endorsements of minority categories of categorical variables) were included. Bonferroni correction for the number of analyses per PRS was used to adjust for multiple comparisons.

**7) PheWAS adjusted by phenotypic externalizing behavior.** Substance use disorders are commonly categorized within the externalizing spectrum of psychiatric disorders, given their overlapping phenotypic and genetic variance^26–28^. SUDs are preceded in many individuals by earlier onset externalizing problems, such as ADHD, oppositional defiant disorder, and conduct disorder symptoms^29^, and all of these phenotypes have shared heritable components ^27,28^. Given the robust associations between SUD PRS and measured externalizing phenotypes in this study, as well as the possibility that genetic risk for SUDs may be expressed earlier as externalizing problems prior to the onset of substance use, we tested whether associations with other phenotypes measured at follow-up are independent of baseline externalizing behaviors by including the baseline CBCL externalizing scale as a covariate analyses for phenotypes exhibiting Bonferroni-correcte significant associations with SUD PRS at follow-up (excluding CBCL externalizing measured at follow-up). Bonferroni correction was employed for all tests within each PRS.

**8) Indirect effects through neuroimaging-derived phenotypes.** In order to explore potential neural mechanisms linking genetic liability to its phenotypic expression, we tested whether baseline imaging-derived phenotypes indirectly linked PRS to two-year follow-up phenotype. First, associations between baseline imaging metrics and two-year follow-up phenotypes were tested. Only imaging metrics and phenotypes that were significantly associated with the same PRS were included, such that associations survived Bonferroni correction in the full sample and were nominally significant (*p*<0.05) in the substance-naive subsample. Linear mixed effects models covarying for baseline age, sex, MRI scanner type, and COVID visit type, and with random intercepts for family and site ID, were conducted. Second, the mediation R package^30^ was used to test whether imaging metrics that were significantly associated with the follow-up phenotype following Bonferroni correction indirectly linked PRS to follow-up phenotypes. Two mixed effects models were run: the first estimated the association between the PRS and brain structure metric (a path), controlling for baseline age, sex, MRI scanner type, the first 10 PCs, and site ID, with a random intercept for family; the second estimated the association between the brain structure metric and the follow-up outcome (b path), controlling for the PRS, follow-up age, sex, MRI scanner type, the first 10 PCs, COVID visit type, and site ID, with a random intercept for family. Finally, the mediate() function was used to compute the indirect effect and standard error via bootstrapping, with 10,000 simulations per model. Site ID was included as a fixed effect, because the mediation package only allows for a single level of nesting. Bonferroni correction was applied to all tests within each PRS.

**9) Cross-ancestry meta-analysis.** The relative lack of statistically significant associations observed in participants most genetically similar to African reference populations (AFR) may be attributable to: a) a true absence of effect between these PRS and the examined phenotypes in this subsample, b) reduced predictive power of the PRS due to smaller discovery GWAS in AFR, and/or c) the smaller target sample size--which, alone or in conjunction with less predictive PRS--could lead to larger standard errors. The small-to-moderate correlations between PheWAS estimates across genetic ancestry groups suggest partially overlapping effects, most of which did not reach statistical significance in AFR.

To derive estimates more generalizable across ancestry groups, we conducted cross-ancestry meta-analyses. For each PRS at both baseline and follow-up, we used the **metafor** R package^31^ to implement random-effects meta-analyses, using the ancestry-stratified effect estimates and standard errors from the primary PheWAS. P-values were Bonferroni-corrected for the (smaller) number of phenotypes analyzed in AFR (940 at baseline; 1,357 at follow-up).

**References**

1.	Ewing SWF, Bjork JM, Luciana M. Implications of the ABCD study for developmental neuroscience. *Dev Cogn Neurosci*. 2018;32:161-164. doi:10.1016/j.dcn.2018.05.003

2.	Martin AR, Gignoux CR, Walters RK, et al. Human Demographic History Impacts Genetic Risk Prediction across Diverse Populations. *Am J Hum Genet*. 2017;100(4):635-649. doi:10.1016/j.ajhg.2017.03.004

3.	Martin AR, Kanai M, Kamatani Y, Okada Y, Neale BM, Daly MJ. Clinical use of current polygenic risk scores may exacerbate health disparities. *Nat Genet*. 2019;51(4):584-591. doi:10.1038/s41588-019-0379-x

4.	Hatoum AS, Colbert SMC, Johnson EC, et al. Multivariate genome-wide association meta-analysis of over 1 million subjects identifies loci underlying multiple substance use disorders. *Nat Ment Health*. 2023;1(3):210-223. doi:10.1038/s44220-023-00034-y

5.	Paul SE, Baranger DAA, Johnson EC, et al. Alcohol milestones and internalizing, externalizing, and executive function: longitudinal and polygenic score associations. *Psychol Med*. 2024;54(10):2644-2657. doi:10.1017/S003329172400076X

6.	Casey BJ, Cannonier T, Conley MI. The Adolescent Brain Cognitive Development (ABCD) study: Imaging acquisition across 21 sites. *Dev Cogn Neurosci*. 2018;32:43-54. doi:10.1016/j.dcn.2018.03.001

7.	Hagler DJ, Hatton S, Cornejo MD, et al. Image processing and analysis methods for the Adolescent Brain Cognitive Development Study. *NeuroImage*. 2019;202:116091. doi:10.1016/j.neuroimage.2019.116091

8.	Ge T, Chen CY, Ni Y, Feng YCA, Smoller JW. Polygenic prediction via Bayesian regression and continuous shrinkage priors. *Nat Commun*. 2019;10(1). doi:10.1038/s41467-019-09718-5

9.	Ruan Y, Lin YF, Feng YCA, et al. Improving polygenic prediction in ancestrally diverse populations. *Nat Genet*. 2022;54(5):573-580. doi:10.1038/s41588-022-01054-7

10.	Chang CC, Chow CC, Tellier LC, Vattikuti S, Purcell SM, Lee JJ. Second-generation PLINK: rising to the challenge of larger and richer datasets. *GigaScience*. 2015;4:7. doi:10.1186/s13742-015-0047-8

11.	Zhou H, Kember RL, Deak JD, et al. Multi-ancestry study of the genetics of problematic alcohol use in over 1 million individuals. *Nat Med*. 2023;29(12):3184-3192. doi:10.1038/s41591-023-02653-5

12.	Toikumo S, Jennings MV, Pham BK, et al. Multi-ancestry meta-analysis of tobacco use disorder identifies 461 potential risk genes and reveals associations with multiple health outcomes. *Nat Hum Behav*. 2024;8(6):1177-1193. doi:10.1038/s41562-024-01851-6

13.	Levey DF, Galimberti M, Deak JD, et al. Multi-ancestry genome-wide association study of cannabis use disorder yields insight into disease biology and public health implications. *Nat Genet*. 2023;55(12):2094-2103. doi:10.1038/s41588-023-01563-z

14.	Deak JD, Zhou H, Galimberti M, et al. Genome-wide association study in individuals of European and African ancestry and multi-trait analysis of opioid use disorder identifies 19 independent genome-wide significant risk loci. *Mol Psychiatry*. 2022;27(10):3970-3979. doi:10.1038/s41380-022-01709-1

15.	Baurley JW, Edlund CK, Pardamean CI, Conti DV, Bergen AW. Smokescreen: a targeted genotyping array for addiction research. *BMC Genomics*. 2016;17:145. doi:10.1186/s12864-016-2495-7

16.	Lam M, Awasthi S, Watson HJ, et al. RICOPILI: Rapid Imputation for COnsortias PIpeLIne. *Bioinformatics*. 2020;36(3):930-933. doi:10.1093/bioinformatics/btz633

17.	Taliun D, Harris DN, Kessler MD. Sequencing of 53,831 diverse genomes from the NHLBI TOPMed Program. *Nature*. 2021;590(7845):290-299. doi:10.1038/s41586-021-03205-y

18.	Bates D, Mächler M, Bolker B, Walker S. Fitting Linear Mixed-Effects Models Using lme4. *J Stat Softw*. 2015;67(1). doi:10.18637/jss.v067.i01

19.	Miller AP, Baranger DAA, Paul SE, et al. Neuroanatomical Variability and Substance Use Initiation in Late Childhood and Early Adolescence. *JAMA Netw Open*. 2024;7(12):e2452027. doi:10.1001/jamanetworkopen.2024.52027

20.	Bartoń K. MuMIn: Multi-model inference, R package version 0.12.0. Published online 2009.

21.	Keller MC. Gene × environment interaction studies have not properly controlled for potential confounders: the problem and the (simple) solution. *Biol Psychiatry*. 2014;75(1):18-24. doi:10.1016/j.biopsych.2013.09.006

22.	Long JA. interactions: Comprehensive, User-Friendly Toolkit for Probing Interactions. Published online February 19, 2019:1.2.0. doi:10.32614/CRAN.package.interactions

23.	Gorelik AJ, Paul SE, Miller AP, et al. Associations between polygenic scores for cognitive and non-cognitive factors of educational attainment and measures of behavior, psychopathology, and neuroimaging in the adolescent brain cognitive development study. *Psychol Med*. 2024;54(13):1-15. doi:10.1017/S0033291724002174

24.	Paul SE, Elsayed NM, Colbert SMC, Bogdan R, Hatoum AS, Barch DM. Family income and polygenic scores are independently but not interactively associated with cognitive performance among youth genetically similar to European reference populations. *Dev Psychopathol*. Published online November 5, 2024:1-15. doi:10.1017/S0954579424001573

25.	Selzam S, Ritchie SJ, Pingault JB, Reynolds CA, O’Reilly PF, Plomin R. Comparing Within- and Between-Family Polygenic Score Prediction. *Am J Hum Genet*. 2019;105(2):351-363. doi:10.1016/j.ajhg.2019.06.006

26.	Kotov R, Krueger RF, Watson D, et al. The Hierarchical Taxonomy of Psychopathology (HiTOP): A Quantitative Nosology Based on Consensus of Evidence. *Annu Rev Clin Psychol*. 2021;17:83-108. doi:10.1146/annurev-clinpsy-081219-093304

27.	Karlsson Linnér R, Mallard TT, Barr PB, et al. Multivariate analysis of 1.5 million people identifies genetic associations with traits related to self-regulation and addiction. *Nat Neurosci*. 2021;24(10):1367-1376. doi:10.1038/s41593-021-00908-3

28.	Kendler KS, Prescott CA, Myers J, Neale MC. The Structure of Genetic and Environmental Risk Factors for Common Psychiatric and Substance Use Disorders in Men and Women. *Arch Gen Psychiatry*. 2003;60(9):929-937. doi:10.1001/archpsyc.60.9.929

29.	Iacono WG, Malone SM, McGue M. Behavioral disinhibition and the development of early-onset addiction: common and specific influences. *Annu Rev Clin Psychol*. 2008;4:325-348. doi:10.1146/annurev.clinpsy.4.022007.141157

30.	Tingley D, Yamamoto T, Hirose K, Keele L, Imai K. mediation: R Package for Causal Mediation Analysis. *J Stat Softw*. 2014;59(5):1-38.

31.	Viechtbauer W. Conducting Meta-Analyses in R with the metafor Package. *J Stat Softw*. 2010;36(3). doi:10.18637/jss.v036.i03
