## supplemental note for "Youth Correlates of Genetic Liability to Substance Use Disorders"

**European-like genetic ancestry	2-6**

**Substance-naive PheWAS	2**

**Simultaneous SUD PRS PheWAS	2-3**

**Phenotypic pruning	3-4**

**PRS-by-time moderation	4**

**Sex moderation	4**

**Within-family PRS effects	5**

**PheWAS adjusted for phenotypic externalizing behavior	5-6**

**African-like genetic ancestry	6-9**

**Substance-naive PheWAS	6**

**Simultaneous SUD PRS PheWAS	6**

**Phenotypic pruning	6**

**PRS-by-time moderation	6**

**Sex moderation	6**

**Within-family PRS effects	7**

**PheWAS adjusted for phenotypic externalizing behavior	7**

**Indirect effects through neuroimaging-derived phenotypes	7**

**Cross-ancestry meta-analysis	7-14**

**References	15**

***European-like Genetic Ancestry***

**Substance-naive PheWAS**. Most (PAU: 96.2%; TUD: 100%; CUD: 88.6%; OUD: 100%; ADDrf: 93.8%) associations between baseline phenotypes and SUD PRS remained significant in the substance-naive sample, following Bonferroni correction for the number of phenotypes (n=10-70; |B|≥0.025, OR≥1.15, *p*≤1.04e-03, *p_Bonferroni_*≤0.048; **Supplemental Table 11**). Associations between PAU PRS and frequency of watching R-rated movies; between CUD PRS and frequency of watching R-rated movies, CBCL externalizing items and DSM-5 depression, past pressured speech, caregiver rulebreaking problems, and total sleep problems; and between ADDrf PRS and caregiver failure to pay debts did not survive correction for multiple tests (|B|≤0.053, OR=1.28, *p*≥7.17e-04, *p_Bonferroni_*≥0.050; **Supplemental Table 11**).

Similarly, the majority (PAU: 84.3%; TUD: 85.1%; CUD: 76.9%; OUD: 97.1%; ADDrf: 84.2%) of associations between follow-up phenotypes and SUD PRS survived Bonferroni correction for the number of phenotypes (n=19-134) in the substance-naive sample (|B|≥0.026, OR≥1.16, *p*≤1.08e-03, *p_Bonferroni_*≤0.048; **Supplemental Table 12**). Associations between PAU PRS and several life events items (e.g., family member arrested, negative impact summary score), youth self-reported total problems and number of psychotic-like experiences, NIH Toolbox picture vocabulary, using one’s phone in the middle of the night, and the number of friends who drink alcohol); TUD PRS and several child behavior and mental health phenotypes (i.e., self-reported hot temper, caregiver-reported destroying one’s own things, suddenly not trusting others, volatile mood, “mania” summary score, and total sleep problems) and fewer sports and activities in the past year; CUD PRS and a number of youth- and caregiver-reported child behavior and mental health phenotypes (e.g., impulsive, aggressive, and internalizing behaviors and traits; bedtime reluctance), caregiver-child conflict, life events (losing close friend, family member with an SUD), lower school involvement (excused absences, disliking school), screentime and media use (weekend time spent editing photos or videos to post on social media, weekday time spent texting, frustration when unable to use screen media), and positive cannabis expectancies (i.e., cannabis helps a person relax); OUD PRS and coffee intake; and ADDrf PRS and coffee intake, having an internet-connected device or TV in one’s bedroom, and frequency of spending time on social media) did not survive correction for multiple tests (|B|≤0.085, OR≤1.25, *p*≥3.91e-04, p_Bonferroni_≥0.051; **Supplemental Table 12**).

**Simultaneous SUD PRS PheWAS.** Unique associations were identified for TUD (baseline: n_Bonf_=2, n_FDR_=21; follow-up: n_Bonf_=3, n_FDR_=34), CUD (baseline: n_Bonf_=20, n_FDR_=136; follow-up: n_Bonf_=21, n_FDR_=128), and OUD PRS (baseline: n_Bonf_=0, n_FDR_=0; follow-up: n_Bonf_=2, n_FDR_=12), but not for PAU PRS, when the four non-ADDrf PRS were entered into the model simultaneously (**Supplemental Tables S13-14)**. CUD PRS was uniquely associated primarily with phenotypes indicating increased impulsivity and externalizing behaviors as measured on the CBCL, K-SADS, youth Brief Problem Monitor, Early Adolescent Temperament Questionnaire, and BIS/BAS. TUD PRS was uniquely linked with increased self-reported time spent on social media at baseline and with difficulty sticking to plans, energy/feelings commonly being at extreme ends, and social jetlag at follow-up. Finally, OUD PRS was independently associated with having more than one social media account and with a higher percent of individuals in one’s neighborhood with a disability (B≤0.032, *p*≤3.52e-05, *p_FDR_*≤0.014, *p_Bonferroni_*≤0.0497).

**Phenotypic Pruning.** Pruning associations based on phenotypic correlation and strength of association (i.e., p-value) with the SUD PRS resulted in a largely domain non-specific monotonic reduction in significant associations. Associations observed to be independent at or below an R^2^ threshold of 0.1 included 50%, 50%, 24%, 40%, and 71% of significant baseline associations for PAU, TUD, CUD, OUD, and ADDrf PRS, respectively, and 39%, 40%, 11%, 34%, and 53% of follow-up associations, respectively (**Supplemental Tables 9-10**). At baseline, these phenotypes included screentime and media use (mature-rated video game frequency, R-rated movie frequency, weekend social media frequency, total weekend screentime), family behavior and mental health (caregiver failure to pay debt, caregiver history of depression, paternal drug problems, caregiver somatic problems, caregiver rule-breaking behavior, caregiver history of anxiety, caregiver history of getting into trouble, and caregiver history of depression), prenatal/postnatal and early development (prenatal tobacco exposure and cigarettes/day before maternal knowledge of pregnancy, unplanned pregnancy, prenatal cannabis exposure before maternal knowledge of pregnancy), substance use and perceptions (total caffeine intake), substance-related environment (cigarette accessibility, different smoking rules in family, different cannabis rules in family), cognition (picture vocabulary, picture sequence memory test, total cognition composite, rey auditory verbal learning test, and list sorting working memory), parent educational attainment, and child behavior and mental health (total hours sleep per night, K-SADS past trauma-related concentration problems and exaggerated startle response, bis/bas fun-seeking, K-SADS depression-related guilt, K-SADS pressured speech, CBCL externalizing problems, CBCL spending time with peers who get into trouble, UPPS-P not getting things done on time, and talking in one’s sleep).

At follow-up, these phenotypes included screentime and media use (weekday social media use, mature-rated video game frequency, R-rated movie frequency, using one’s phone in the middle of the night, caregiver worry that youth will post something inappropriate online, total weekday screentime, editing photos or videos to post on social media, only screen media seeming to help when having a bad day, watching/streaming movies/TV, having at least one social media account, having an internet-connected device or TV in one’s bedroom), life events (new stepparent, family member having an SUD, sum of life events that affected the youth), substance use and perceptions (perceived smokeless tobacco use harm, perceived e-cigarette harm, expecting cannabis to be relaxing, energy drink intake, caffeine summary score), substance-related environment (number of friends who use “other” tobacco products), cognition (picture vocabulary), family environment (parent education, partner not youth’s biological parent, caregiver-child conflict due to school problems, less frequent eating together as a family), peer environment (discrimination due to one’s ethnicity, problems with bullying, involvement with peers who break rules, peers suspended from school, victim of gossip), school-related (detention/suspension, excused absences, drop in grades, disliking school, lower school environment), neighborhood/external environment (discrimination due to one’s sexuality, discrimination due to one’s weight, lower neighborhood high-school employment), physical health (past year number of sports and related activities), and child behavior and mental health (BIS/BAS going for something right away when one sees an opportunity to get something they want, self-reported disobedience at school, self-reported total problems, caregiver-reported destroying one’s own things, CBCL DSM-5 conduct disorder, EATQ being scared of the dark at night, EATQ struggling to stick to a plan, EATQ lower inhibitory, one’s feelings/energy usually up or down but rarely in the middle, social jetlag, number of times awake during the night, severity of psychotic-like experiences, suddenly not trusting others when seem to be watching/talking about them, number of hours reading per week for pleasure, number of hours’ sleep per night, UPPS likes new/thrilling things even if they are a little scary).

**PRS-by-time moderation.** No PRS was significantly associated with change in any phenotype measured at both assessment waves, as indicated by a lack of significant wave-x-PRS interactions following Bonferroni or FDR correction (all |B|≤0.37, all *p*≥9.84e-04, all *p_FDR_*≥0.45, all *p_Bonferroni_*≥0.79; **Supplemental Table 15**).

**Sex Moderation.** The significant interaction between PAU PRS and sex in predicting baseline ksads_9_37_p (caregiver-reported present active avoidance of phobic object; B_PRS_=0.31, B_PRS_*sex=-0.43, *p_PRS_***sex*=3.77e-05, *p_FDR PRS_**sex=0.032, *p_Bonferroni PRS_***sex*=0.038) involved a significant simple slope for female (B=0.31, *p*=5.12e-5) but not male participants (B=-0.12, *p*=-0.09). A Johnson-Neyman test indicated that the interaction was significant outside of the PAU PRS range -0.54 to 0.54 (**Supplemental Table S16; Extended Data Figure 4a**).

The significant interaction between CUD PRS and baseline pps_2_bother_yn (bothered by hearing strange sounds; B_PRS_=0.40, B_PRS_*sex=-0.66, *p_PRS_*sex*=1.45e-05, *p_FDR PRS_*sex*=0.015, *p_Bonferroni PRS_*sex*=0.015) involved statistically significant simple slopes for female (B=0.41, *p*=2.16e-04) and male (B=-0.24, *p*=0.02) participants that were in the opposite direction. The Johnson Neyman interval for which the interaction was significant was outside of the CUD PRS range of -0.45 to 0.48 (**Supplemental Table S16; Supplemental Figure 4b**).

The significant interaction between TUD PRS and follow-up sleepdisturb5_p (feeling anxious when falling asleep; B_PRS_=0.10, B_PRS_*sex=-0.12, *p_PRS_*sex*=1.45e-05, *p_FDR PRS_*sex*=0.015, *p_Bonferroni PRS_*sex*=0.015) involved a statistically significant simple slope for female (B=0.10, *p*=5.63e-06) but not male (B=-0.02, *p*=0.41) participants. The Johnson Neyman interval for which the interaction was significant was outside of the TUD PRS range of -2.41 to -0.57 (**Supplemental Table S17; Extended Data Figure 4c**).

The significant interaction between OUD PRS and follow-up resiliency6b_y (number of close friends that are girls; B_PRS_=-0.019, B_PRS_*sex=0.10, *p_PRS_*sex*=3.13e-05, *p_FDR PRS_*sex*=0,043, *p_Bonferroni PRS_*sex*=0.043) involved a statistically significant simple slope for male (B=0.08, *f*=3.22e-07) but not female (B=-0.02, *p*=0.26) participants. The Johnson Neyman interval for which the interaction was significant was outside of the range of observed data (**Supplemental Table S17; Extended Data Figure 4d**).

**Within-family PRS effects.** Following Bonferroni correction for the number of tests, within-family PRS effects were identified for PAU (baseline, n=1; follow-up, n=0), CUD (baseline, n=0; follow-up, n=5), and TUD (baseline, n=1; follow-up, n=1) PRS. Within-family PAU PRS was associated with increased caffeinated soda consumption (B=0.063, *p*=5.38e-05, *p_FDR_*=1.29e-03, *p_Bonferroni_*=1.29e-03) at baseline. Within-family TUD PRS was associated with increased caregiver self-reported restlessness at baseline (B=0.028, *p=*1.55e-03, *p_FDR_*=0.028, *p_Bonferroni_*=0.045) and increased caregiver-reported tendency for youth’s moods to be generally “up or down, but rarely in the middle” on the General Behavior Inventory - Mania at two-year follow-up (B=0.055, *p=*7.82e-06, *p_FDR_*=5.16e-04, *p_Bonferroni_*=5.16e-04). Finally, within-family CUD PRS was associated with increased CBCL temper tantrums and CBCL breaking rules at school, home, or elsewhere; caregiver-child conflict; problems with bullying; and youth self-reported arguing (B≥0.075, *p*≤2.45e-04, *p_FDR_*≤6.41e-03, *p_Bonferroni_*≤0.032; **Supplemental Tables 18-19; Extended Data Figure 5**).

**PheWAS adjusted by phenotypic externalizing behavior.** The majority (PAU: 100%; TUD: 92.5%; CUD: 58.5%; OUD: 100%; ADDrf 100%) of Bonferroni-significant associations between SUD PRS and follow-up phenotypes remained significant after covarying for baseline CBCL externalizing problems and correcting for the number of tests (the number of Bonferroni-significant primary PheWAS associations, except CBCL externalizing problems) within each PRS (**Supplemental Table 20**).

The five phenotypes no longer associated with TUD PRS were related to externalizing (i.e., CBCL DSM-5 conduct problems and rulebreaking behavior, EATQ aggression and problems with inhibition) or sleep problems (i.e., total sleep disturbances score) and were not particularly strongly associated with TUD PRS in primary analyses (i.e., *p_Bonferroni_*≥0.014). Social jetlag remained the strongest association, and phenotypes related to screentime and media use, caffeine intake, and school performance also continued to exhibit strong associations (**Supplemental Table 20**).

Of the 56 phenotypes no longer significantly associated with CUD PRS, 22 came from the CBCL and 11 from the EATQ (compared to four and two, respectively, that remained associated); many of these reflected externalizing, impulsive, and/or inattentive behavior. Most screentime and media use items continued to be associated with CUD PRS (i.e., 13 phenotypes representing time spent on various screen-related activities, having social media, and caregivers following their children on social media, compared to three phenotypes no longer associated with CUD PRS that reflected caregivers’ reports of concern about, and motivations for, their child’s screen media use). The majority of youth self-report of behavior problems, school performance, social jetlag, stressful life events, psychotic-like events, school involvement, and caffeine intake phenotypes remained significantly associated with CUD PRS following adjustment for baseline CBCL externalizing problems and Bonferroni correction ((**Supplemental Table 20**).

***African-like Genetic Ancestry***

**Substance-naive PheWAS**. Though there were no significant associations observed in the primary PheWAS at baseline, results from the substance-naive sample are shown in **Supplemental Table 23**. All follow-up phenotypes associated with AUD and CUD PRS in primary analyses remained significantly associated with the PRS in the substance-naive subsample following Bonferroni correction for the number of tests (one and six, respectively; |B|≥0.11, *p*≤3.92e-04, *p_Bonferroni_*≤2.35e-03; **Supplemental Table 24**).

**Simultaneous SUD PRS PheWAS.** No SUD PRS was independently associated with any phenotype at baseline when AUD, TUD, CUD, and OUD PRS were entered simultaneously into the model (|B|≤0.25, OR≤1.52, *p*≥9.37e-05, *p_FDR_*≥0.088, *p_Bonferroni_*≥0.088; **Supplemental Table 25**). CUD PRS was significantly independently associated with CBCL DSM-5 ADHD, attention problems, and disobedience at home (B≥0.15, *p*≤1.52e-05, *p*_FDR_≤6.88e-03, *p*_Bonferroni_≤0.021; **Supplemental Table 26**).

**Phenotypic Pruning.** Pruning associations with CUD PRS at follow-up based on phenotypic correlation and strength of association (i.e., p-value) showed that CBCL externalizing problems and DSM-5 ADHD had the strongest and most independent associations (R^2^=0.6), followed by CBCL DSM-5 conduct disorder and total problems (R^2^=0.8), CBCL attention problems (R^2^=0.9), and aggressive problems (pruned immediately at R^2^=1.0; **Supplemental Table 22**).

**PRS-by-time moderation.** No PRS was significantly associated with change in any phenotype measured at both assessment waves (all interaction |B|≤0.37, all p≥1.16e-03, all *p*_FDR_≥0.51, all *p*_Bonferroni_≥0.72; **Supplemental Table 27**).

**Sex Moderation.** No significant PRS*sex indirections were identified following Bonferroni correction (all |B|≤0.67, all *p*≥1.72e-04, all *p*_FDR_≥0.16, all *p*_Bonferroni_≥0.19; **Supplemental Tables 28-29).**

**Within-family PRS effects.** The single Bonferroni-significant AUD PRS association at follow-up was not significant at the within-family level (B=0.015, *p*=0.78). Within-family CUD PRS was positively associated with all six CBCL scales following FDR correction for six tests, and the association with the CBCL DSM-5 conduct disorder scale additionally survived Bonferroni correction (B≥0.071, *p*≤0.030, *p_FDR_*≤0.036, *p_Bonferroni_*≥0.048; **Supplemental Table 30**).

**PheWAS adjusted by phenotypic externalizing behavior.** AUD PRS remained significantly negatively associated with knowing how to get in touch with caregivers when they are not at home after covarying for baseline CBCL externalizing problems (B=-0.12, *p*=2.77e-05). CUD PRS remained significantly positively associated with the CBCL DSM-5 conduct disorder scale (B=0.054, *p*=9.88e-03, *p*_Bonferroni_=0.049) but not with the CBCL DSM-5 ADHD scale or CBCL aggressive, attention, or total problems (B≤0.050, *p*≥0.063, *p*_Bonferroni_≥0.31; **Supplemental Table 31**).

***Indirect effects through neuroimaging-derived phenotypes.*** The brain-wide and phenome-wide significant imaging associations in individuals of European ancestry that were also observed (*p*<0.05) in the substance-naive sample were carried forward into indirect effects analyses. Following Bonferroni correction for the number of tests within each PRS (PAU PRS: 51 tests; TUD PRS: 67*3 brain regions = 201 tests; CUD PRS: 134*2 = 268 tests), 10, 5, and 11 associations between imaging metric and follow-up phenotype were identified for PAU, TUD, and CUD PRS, respectively (Supplemental Table 2.6.32). With the exception of RSFC-sensorimotor hand-sensorimotor mouth network indirectly linking PAU PRS to one’s caregiver’s partner *not* being a biological parent (|average causal mediation effect| = 5.04e-04, proportion mediated = 0.027, *p*=5.60e-03, *p_Bonferron_*=0.056), all of these imaging metrics significantly indirectly linked the PRS to the respective follow-up phenotype following Bonferroni correction for all tests within each PRS (|average causal mediation effect| ≥ 1.34e-03, proportion mediated ≥ 0.0275, *p*≤5.40e-03, *p_Bonferroni_*≤0.027^[1]^; **Supplemental Table 35; Extended Data Figures 8-10**).

***Cross-ancestry meta-analysis.***

**Baseline** (see **Supplemental Table 36**).

***Problematic Alcohol Use/Alcohol Use Disorder PRS*.** Of the 25 phenotypes measured in both genetic ancestry groups that were Bonferroni-significant in EUR, 7 were Bonferroni-significant in the cross-ancestry meta-analysis: Greater weekend social media hours, mature-rated video game frequency, R-rated movie frequency, total weekend screentime, prenatal cigarettes per day both before and after pregnancy knowledge, and unplanned pregnancy (Meta |B|≥0.023, OR=1.18, *p*≤2.96e-05, *p_Bonferroni_*≤0.028; EUR |B|≥0.023, OR=1.21, *p*≤2.63e-05, *p_Bonferroni_*≤0.033; AFR |B|≤0.021, OR=1.11, *p*≥5.67e-03, *p_FDR_*≥0.92). An additional 3 phenotypes were Bonferroni-significant in EUR and FDR-significant in the meta-analysis: fewer hours of sleep per night, caregiver history of alcohol problems, and paternal history of alcohol problems (Meta B=0.059, ORs=1.28, *p_FDR_*≤0.021, *p_Bonferroni_*≥0.065; EUR B=0.089, OR≥1.35, *p*≤3.50e-05, *p_Bonferroni_*≤0.045; AFR B=0.037, OR≤1.17, *p*≥0.089, *p_FDR_*≥0.92). Four phenotypes were FDR-significant in EUR and Bonferroni-significant in the meta-analysis: BIS/BAS doing things on the spur of the moment, CBCL DSM-5 conduct disorder, lower caregiver educational attainment, and CBCL swearing/obscene language (Meta |B|≥0.024, *p*≤4.15e-03, *p_Bonferroni_*≤0.046; EUR |B|≥0.026, *p*≤1.25e-04, *p_FDR_*≤4.27e-03, *p_Bonferroni_*≥0.066). Q-tests indicated that the ancestry-specific effect sizes for these phenotypes were not significantly heterogeneous (Q≤1.50*,* Q*_Pval_*≥0.22) and that percent of the total variance attributed to heterogeneity (i.e., I^2^) was between 0 and 33.3%. For the 11 phenotypes Bonferroni-significant in EUR but not significant following multiple testing correction in the meta-analysis, the Q-tests were largely significant (10 Q*_Pval_*≤0.049, 1 Q*_Pval_*=0.21), and I^2^=36.9-91.5. These metrics of heterogeneity should be interpreted with caution, however, because only two effect sizes are contributing to each meta-regression; the Q-test may be underpowered, and the I^2^ statistics may be biased in either direction^1,2^.

***Tobacco Use Disorder PRS*.** Of the 30 phenotypes measured in both genetic ancestry groups that were Bonferroni-significant in EUR, 9 were Bonferroni-significant in the cross-ancestry meta-analysis: Prenatal tobacco exposure and cigarettes/day pre pregnancy knowledge and prenatal tobacco exposure after pregnancy knowledge, caregiver self-reported failure to pay debts/other financial obligations, lower NIH Toolbox list sorting working memory, fewer hours of sleep per night, BIS/BAS fun-seeking, CBCL lying/cheating, and CBCL DSM-5 conduct disorder (Meta |B|≥0.029, OR≥1.41, *p*≤2.26e-05, *p_Bonferroni_*≤0.021; EUR |B|≥0.030, OR≥1.46, *p*≤3.25e-05, *p_Bonferroni_*≤0.041; AFR |B|≤0.076, OR≤1.37, *p*≥8.74e-04, *p_FDR_*≥0.61). An additional 7 phenotypes were FDR-significant in EUR and Bonferroni-significant in the meta-analysis: Prenatal cannabis exposure pre pregnancy knowledge, maternal history of depression, past-year detention or suspension, greater cigarette accessibility, greater UPPS-P positive urgency, and greater CBCL rule-breaking and externalizing problems (Meta |B|≥0.031, OR≥1.18, *p*≤3.04e-03, *p_Bonferroni_*≤0.049; EUR |B|≥0.033, OR≥1.18, *p*≤1.42e-03, *p_FDR_*≤0.017, *p_Bonferroni_*≥0.13; AFR |B|≤0.059, OR≤1.28, *p*≥8.77e-03, *p_FDR_*≥0.61). Q-tests indicated that the ancestry-specific effect sizes for these phenotypes were not significantly heterogeneous (Q≤1.61*,* Q*_Pval_*≥0.20) and that percent of the total variance attributed to heterogeneity (i.e., I^2^) was between 0 and 37.9%. For the 21 phenotypes Bonferroni-significant in EUR but not significant following multiple testing correction in the meta-analysis, the Q-tests were largely significant (15 Q*_Pval_*≤0.030, 6 Q*_Pval_*=0.057-0.10), and I^2^=62.4-94.4; but should be interpreted with caution.

***Cannabis Use Disorder PRS*.** Of the 70 phenotypes measured in both genetic ancestry groups that were Bonferroni-significant in EUR, 45 were Bonferroni-significant in the cross-ancestry meta-analysis. These included CBCL scales (externalizing, total problems, aggressive, attention problems, social problems, DSM-5 conduct disorder, DSM-5 oppositional defiant disorder, DSM-5 ADHD, DSM-5 depressive disorder, and the stress scale) and 12/15 CBCL items, 12/13 KSADS ADHD and ODD items, UPPS-P difficulty getting things done on time, the PGMI mania summary score, prenatal tobacco and cannabis exposure pre pregnancy knowledge (but not unplanned pregnancy), greater total sleep problems (but not hours per day), greater cigarette accessibility, and caregiver self-reported rule-breaking (Meta |B|≥0.032, OR≥1.17, *p*≤3.88e-05, *p_Bonferroni_*≤0.037; EUR |B|≥0.030, OR≥1.17, *p*≤3.36e-05, *p_Bonferroni_*≤0.043; AFR |B|≤0.10, OR≤1.28, *p*≥1.49e-04, *p_FDR_*≥0.063). An additional 8 phenotypes were Bonferroni-significant in EUR and FDR-significant in the meta-analysis: 2 CBCL items (demands attention, disobedient in school), teacher-rated difficulties concentrating, disorders of initiating and maintaining sleep, lower school performance, caregiver history of depression and paternal history of drug problems, and lower caregiver education (Meta |B|≥0.033, OR≥1.27, *p*≤9.96e-03, *p_FDR_*≤0.041, *p_Bonferroni_*≥0.12; EUR |B|≥0.034, OR≥1.36, *p*≤1.43e-05, *p_Bonferroni_*≤0.018; AFR |B|≤0.069, OR≤1.18, *p*≥9.86e-03, *p_FDR_*≥0.18). Twenty five phenotypes were FDR-significant in EUR and Bonferroni-significant in the meta-analysis: CBCL thought problems and 5 CBCL items related to externalizing and attention problems, 11 KSADS items related to ADHD and ODD, K-SADS past distractibility on the Bipolar Disorders module, greater severity of psychotic-like experiences, 2 teacher-reported items related externalizing and attention problems, prenatal cannabis exposure quantity pre pregnancy knowledge, caregiver-child conflict and conflict related to youth’s moods, and poorer grades in school (Meta |B|≥0.019, OR≥1.13, *p*≤5.24e-05, *p_Bonferroni_*≤0.0489; EUR |B|≥0.018, OR≥1.13 *p*≤1.38e-03, *p_FDR_*≤9.11e-03, *p_Bonferroni_*≥0.063; AFR |B|≤0.088, OR≤1.28, *p*≥2.04e-03, *p_FDR_*≥0.11). Q-tests indicated that the ancestry-specific effect sizes for these phenotypes were not significantly heterogeneous (Q≤1.87*,* Q*_Pval_*≥0.17) and that percent of the total variance attributed to heterogeneity (i.e., I^2^) was between 0 and 46.6%. For the 19 phenotypes Bonferroni-significant in EUR but not significant following multiple testing correction in the meta-analysis, the Q-tests were largely significant (13 Q*_Pval_*≤0.041, 6 Q*_Pval_*=0.050-0.13), and I^2^=56.1-90.9; but should be interpreted with caution.

***Opioid Use Disorder PRS***. Of the 10 phenotypes measured in both genetic ancestry groups that were Bonferroni-significant in EUR, 2 were Bonferroni-significant in the cross-ancestry meta-analysis: Caffeine intake summary score and caffeinated soda intake (Meta B≥0.047, *p*≤4.79e-06, *p_Bonferroni_*≤4.50e-03; EUR B≥0.050, *p*≤1.62e-05, *p_Bonferroni_*≤0.021; AFR B≤0.054, *p*≥0.017, *p_FDR_*≥0.99). An additional 2 phenotypes were Bonferroni-significant in EUR and FDR-significant in the meta-analysis: NIH Toolbox picture sequence memory test and picture vocabulary test (Meta B≤-0.049, *p*≤8.31e-04, *p_FDR_*≤0.049, *p_Bonferroni_*≥0.17; EUR B≤-0.059, *p*≤8.42e-06, *p_Bonferroni_*≤0.011; AFR |B|≥0.026, *p*≥0.25). Prenatal tobacco exposure pre pregnancy knowledge was further Bonferroni-significant in the meta-analysis and FDR-significant in EUR (Meta OR=1.25, *p*=2.02e-05, *p_Bonferroni_*=0.019; EUR OR=1.28, *p*=2.26e-04, *p_FDR_*=0.012, *p_Bonferroni_*=0.29; AFR OR=1.20, *p*=0.025, *p_FDR_*=0.99). Q-tests indicated that the ancestry-specific effect sizes for these phenotypes were not significantly heterogeneous (Q≤1.34*,* Q*_Pval_*≥0.25) and that percent of the total variance attributed to heterogeneity (i.e., I^2^) was between 0 and 25.6%. For the 6 phenotypes Bonferroni-significant in EUR but not significant following multiple testing correction in the meta-analysis, the Q-tests were largely significant (5 Q*_Pval_*≤0.026, 1 Q*_Pval_*=0.081), and I^2^=67.2-92.7; but should be interpreted with caution.

***Addiction Risk Factor PRS***. Of the 16 phenotypes measured in both genetic ancestry groups that were Bonferroni-significant in EUR, 5 were Bonferroni-significant in the cross-ancestry meta-analysis: BIS/BAS fun-seeking and doing things in the spur of the moment, total weekend screentime and R-rated movie frequency, and prenatal cigarettes/day pre pregnancy knowledge (Meta B≥0.028, *p*≤3.02e-05, *p_Bonferroni_*≤0.028; EUR B≥0.032, *p*≤3.65e-05, *p_Bonferroni_*≤0.046; AFR B≤0.049, *p*≥0.054). One phenotype (unplanned pregnancy) was additionally Bonferroni-significant in EUR and FDR significant in the meta-analysis (Meta OR=1.17, *p*=1.18e-03, *p_FDR_*=0.030, *p_Bonferroni_*=1.00; EUR OR=1.21, *p*=1.95e-07, *p_Bonferroni_*=2.47e-04; AFR OR=1.10, *p*=0.17). Q-tests indicated that the ancestry-specific effect sizes for these phenotypes were not significantly heterogeneous (Q≤1.77*,* Q*_Pval_*≥0.18) and that percent of the total variance attributed to heterogeneity (i.e., I^2^) was between 0 and 43.5%. For the 10 phenotypes Bonferroni-significant in EUR but not significant following multiple testing correction in the meta-analysis, 3 Q-tests were significant (Q*_Pval_*≤0.038) and 7 were not (Q*_Pval_*=0.098-0.23), and I^2^=30.7-87.7; but should be interpreted with caution.

**Follow-up** (see **Supplemental Table 37**).

***Problematic Alcohol Use/Alcohol Use Disorder PRS*.** Of the 50 phenotypes measured in both genetic ancestry groups that were Bonferroni-significant in EUR, 9 were Bonferroni-significant in the cross-ancestry meta-analysis: Total “bad” life events, the summary score of the degree to which “bad” life events affected the youth, CBCL DSM-5 conduct disorder and rule-breaking behavior, youth-reported total problems, psychotic-like experiences severity score, social media frequency, using one’s phone/device in the middle of the night, and spending time with peers who have been suspended (Meta B≥0.044, *p*≤2.25e-05, *p_Bonferroni_*≤0.031; EUR B≥0.047, *p*≤1.32e-05, *p_Bonferroni_*≤0.022; AFR B≤0.068, *p*≥0.011, *p_Bonferroni_*=1.00). An additional 5 phenotypes were Bonferroni-significant in EUR and FDR-significant in the meta-analysis: Total number of life events and the summary score of the degree to which the youth was affected by life events, lower scores on the NIH Toolbox Picture Vocabulary Test, UPPS-P liking new, thrilling things, and greater weekend social media hours (Meta |B|≥0.050, *p*≤2.96e-03, *p_FDR_*≤0.045, *p_Bonferroni_*≥0.077; EUR |B|≥0.060, *p*≤2.91e-05, *p_Bonferroni_*≤0.049; AFR |B|≤0.039, *p*≥0.16). Seven phenotypes were FDR-significant in EUR and Bonferroni-significant in the meta-analysis: Youth-reported externalizing and attention problems, lower EAT attention and effortful control, CBCL destroying one’s own things, not getting along with one’s teachers, and being chased by another youth as though they were trying to hurt them (Meta |B|≥0.026, *p*≤3.66e-05, *p_Bonferroni_*≤0.0497; EUR |B|≥0.027, *p*≤3.09e-04, *p_FDR_*≤5.58e-03, *p_Bonferroni_*≥0.062; AFR |B|≤0.061, *p*≥0.042, *p_FDR_*≥0.87). Q-tests indicated that the ancestry-specific effect sizes for these phenotypes were not significantly heterogeneous (Q≤1.80*,* Q*_Pval_*≥0.18) and that percent of the total variance attributed to heterogeneity (i.e., I^2^) was between 0 and 44.5%. For the 36 phenotypes Bonferroni-significant in EUR but not significant following multiple testing correction in the meta-analysis, the majority of Q-tests were significant (23 Q*_Pval_*≤0.048, 13 Q*_Pval_*=0.056-0.21), and I^2^=35.6-93.0, but should be interpreted with caution.

***Tobacco Use Disorder PRS*.** Of the 67 phenotypes measured in both genetic ancestry groups that were Bonferroni-significant in EUR, 23 were Bonferroni-significant in the cross-ancestry meta-analysis. These included CBCL externalizing scales and items (externalizing, DSM-5 conduct disorder, rule-breaking problems, destroying one’s own things, and lying/cheating); youth-reported attention problems and having a hot temper, and teacher-reported total problems; EATQ scales and items (aggression, attention, putting off working, slamming the door when angry, and being rude), total sleep problems, poorer grades in school, spending time with peers who have been suspended, having experienced reputational victimization, social media frequency, having a family member with an SUD, coffee consumption, and lower perceived harm of regular e-cigarette and regular smokeless tobacco use (Meta |B|≥0.031; OR=1.23, *p*≤8.87e-06, *p_Bonferroni_*≤0.012; EUR |B|≥0.033, OR=1.25, *p*≤2.63e-05, *p_Bonferroni_*≤0.045; AFR |B|≤0.076, OR=1.17, *p*≥0.010, *p_FDR_*≥0.53). An additional 5 phenotypes were Bonferroni-significant in EUR and FDR-significant in the meta-analysis: The summary score of the degree to which the youth was affected by life events, lower EATQ effortful control, creator caffeine summary score, mature-rated video game frequency, and having experienced detention or suspension (Meta |B|≥0.057, OR=1.29, *p*≤3.32e-03, *p_FDR_*≤0.031, *p_Bonferroni_*≥0.16; EUR |B|≥0.067, OR=1.37, *p*≤4.14e-06, *p_Bonferroni_*≤7.03e-03; |B|≤0.036, OR=1.20, *p*≥0.028, *p_FDR_*≥0.64). Nine phenotypes were FDR-significant in EUR and Bonferroni-significant in the meta-analysis: CBCL aggressive problems and lacking guilt after misbehaving and having sudden changes in one’s mood, youth-reported trouble concentrating and sitting still, EATQ greater negative affect and getting sidetracked, UPPS-P acting without thinking when happy, and being the victim of rumors (Meta |B|≥0.034, *p*≤3.63e-05, *p_Bonferroni_*≤0.049; EUR |B|≥0.032, *p*≤2.99e-04, *p_FDR_*≤4.14e-03, *p_Bonferroni_*≥0.056; AFR |B|≤0.068, *p*≥0.013, *p_FDR_*≥0.54). Q-tests indicated that the ancestry-specific effect sizes for these phenotypes were not significantly heterogeneous (Q≤2.17*,* Q*_Pval_*≥0.14) and that percent of the total variance attributed to heterogeneity (i.e., I^2^) was between 0 and 53.9%. For the 39 phenotypes Bonferroni-significant in EUR but not significant following multiple testing correction in the meta-analysis, there was evidence of heterogeneity in effect sizes (12 Q*_Pval_*≤0.041, 27 Q*_Pval_*=0.053-0.21; I^2^=35.4-94.1; but these should be interpreted with caution).

***Cannabis Use Disorder PRS*.** Of the 133 phenotypes measured in both genetic ancestry groups that were Bonferroni-significant in EUR, 58 were Bonferroni-significant in the cross-ancestry meta-analysis. These included higher CBCL externalizing, attention, rule-breaking, and aggressive problems, CBCL DSM-5 oppositional defiant and conduct disorder scales, and 14 individual CBCL items loading onto these scales; lower EATQ effortful control, inhibitory control, attention, and activation scales, and 8 EATQ items reflecting distractibility, impulsivity, irritability, and procrastination; greater UPPS-P positive and negative urgency and two UPPS-P items; BIS/BAS going for what one wants right away; youth self-reported tendency to argue a lot and feeling worthless; teacher-reported failure to finish things once started; caregiver-reported general behavior inventory summary score and items reflecting rapid mood shifts and irritability alongside happiness and energy; total number and severity of psychotic-like experiences; greater total sleep problems, disorders of initiating and maintaining sleep, and fewer total hours of sleep per night; concentrating too much on parts over the whole; caregiver-youth conflict and conflict related to moods; lower school performance; caregiver self-reported rule-breaking; overt aggression; and having a family member with an SUD (Meta |B|≥0.033, OR≥1.19, *p*≤2.81e-05; *p_Bonferroni_*≤0.0381; EUR |B|≥0.035, OR≥1.18, *p*≤2.74e-05, *p_Bonferroni_*≤0.047). Or these, the CBCL externalizing, aggressive, and DSM-5 conduct disorder scales were also significantly associated with CUD_PRS_ in AFR following Bonferroni correction (B≥0.12, *p*≤2.72e-05, *p_Bonferroni_*≤0.037, and CBCL DSM-5 oppositional defiant disorder and rule-breaking scales and an item about rule-breaking were significant following FDR correction (B≥0.14, *p*≤2.15e-04, *p_FDR_*≤0.43, *p_Bonferroni_*≥0.19; all other |B|≤0.11, OR≤1.24, *p*≥1.01e-03, *p_FDR_*≥0.077). Three phenotypes were Bonferroni-significant in EUR and AFR and FDR-significant in the meta-analysis: CBCL DSM-5 ADHD, attention problems, and total problems (Meta B≥0.11, *p*≤4.91e-03, *p_FDR_*≤0.029, *p_Bonferroni_*≥0.097; EUR B≥0.083, *p*≤2.30e-07, *p_Bonferroni_*≤3.90e-04; AFR B≥0.14, *p*≤3.44e-05, *p_Bonferroni_*≤0.047). Two associations were Bonferoni-significant in EUR and FDR-significant in AFR and the meta-analysis: higher CBCL stress scale score and disobedience in one’s home (Meta B≥0.091, *p*≤2.25e-03, *p_FDR_*≤0.015, *p_Bonferroni_*≥0.082; EUR B≥0.068, *p*≤1.64e-07, *p_Bonferroni_*≤0.041; AFR B≥0.13, *p*≤2.14e-04, *p_FDR_*≤0.024, *p_Bonferroni_*≥0.059). Ten associations were Bonferroni-significant in EUR, FDR-significant in the meta-analysis, and not significant following multiple testing correction in AFR, including two CBCL items reflecting externalizing behavior; youth-reported attention problems; poor school grades and having been suspended or sent to detention; spending time with peers who have been suspended; and the total number of bad life events (Meta B≥0.036, OR≥1.20, *p*≤4.79e-03, *p_FDR_*≤0.029, *p_Bonferroni_*≥0.075; EUR B≥0.031, OR≥1.27, *p*≤2.71e-06, *p_Bonferroni_*≤4.61e-03; AFR B≤0.053, OR≤1.22, *p*≥1.25e-03, *p_FDR_*≥0.077). Twenty phenotypes demonstrated Bonferroni-significant associations in the meta-analysis and FDR-significant associations in EUR, but non-significant associations following multiple testing correction in AFR (Meta B≥0.032, OR≥1.17, *p*≤3.35e-05, *p_Bonferroni_*≤0.046; EUR B≥0.029, OR≥1.16 , *p*≤1.49e-03, *p_FDR_*≤8.32e-03, *p_Bonferroni_*≥0.053; AFR B≤0.099, OR≤1.43, *p*≥1.64e-03, *p_FDR_*≥0.089). Q-tests indicated that the ancestry-specific effect sizes for these phenotypes were not significantly heterogeneous (92 Q≤2.94*,* Q*_Pval_*≥0.087; 1 Q=4.88, Q*_Pval_*=0.027) and that percent of the total variance attributed to heterogeneity (i.e., I^2^) was between 0 and 79.5%. For the 60 phenotypes Bonferroni-significant in EUR but not significant following multiple testing correction in the meta-analysis, there was evidence of heterogeneity in effect sizes (48 Q*_Pval_*≤0.048, 12 Q*_Pval_*=0.050-0.16; I^2^=48.8-95.0; but these should be interpreted with caution).

***Opioid Use Disorder PRS*.** Of the 35 phenotypes measured in both genetic ancestry groups that were Bonferroni-significant in EUR, two were Bonferroni-significant in the cross-ancestry meta-analysis: Youth self-reported tendency to argue a lot, and frequency of talking or texting on the phone (Meta B≥0.060, *p*≤1.56e-05, *p_Bonferroni_*≤0.021; EUR B≥0.066, *p*≤2.85e-06, *p_Bonferroni_*≤4.84e-03; AFR B≤0.034, *p*≥0.24). An additional 5 phenotypes were Bonferroni-significant in EUR and FDR-significant in the meta-analysis: Greater caffeine use summary score, greater neighborhood socioeconomic social vulnerability scores, lower neighborhood social and economic child opportunity index scores, lower scores on the NIH Toolbox picture vocabulary test, and lower perceived harm for regular smokeless tobacco use (Meta |B|≥0.054, *p*≤5.85e-04, *p_FDR_*≤0.038, *p_Bonferroni_*≥0.091; EUR |B|≥0.063, *p*≤2.24e-05, *p_Bonferroni_*≤0.038; AFR |B|≤0.046, *p*≥0.17). Five phenotypes were significant following Bonferroni correction in the meta-analysis and FDR correction in EUR: lower neighborhood per capita income, greater neighborhood child opportunity index housing public assistance, lower neighborhood child opportunity index economic resource index, and greater caffeinated soda intake (Meta |B|≥0.047, *p*≤2.58e-05, *p_Bonferroni_*≤0.035; EUR |B|≥0.047, *p*≤2.93e-04, *p_FDR_*≤6.99e-03, *p_Bonferroni_*≥0.085; AFR |B|≤0.074, *p*≥0.015, *p_FDR_*≥0.996). Q-tests indicated that the ancestry-specific effect sizes for these phenotypes were not significantly heterogeneous (Q≤1.36*,* Q*_Pval_*≥0.24) and that percent of the total variance attributed to heterogeneity (i.e., I^2^) was between 0 and 26.5%. For the 28 phenotypes Bonferroni-significant in EUR but not significant following multiple testing correction in the meta-analysis, there was some evidence of heterogeneity in effect sizes (13 Q*_Pval_*≤0.045, 15 Q*_Pval_*=0.073-0.24; I^2^=26.9-89.5; but these should be interpreted with caution).

***Addiction Risk Factor PRS*.** Of the 18 phenotypes measured in both genetic ancestry groups that were Bonferroni-significant in EUR, eight were Bonferroni-significant in the cross-ancestry meta-analysis: Greater maximum caffeine intake, total number of life events, sum of life events that affected the youth, total number of bad life events, sum of life events that affected youth in a bad way, teacher-reported school disobedience, social media use frequency, and having a TV or internet in one’s bedroom (Meta B≥0.043, OR=1.15, *p*≤8.95e-06, *p_Bonferroni_*≤0.012; EUR B≥0.043, OR=1.16, *p*≤1.78e-05, *p_Bonferroni_*≤0.030; AFR B≤0.071, OR=1.11, *p*≥0.011, *p_FDR_*≥0.20). Seven phenotypes were significant following Bonferroni correction in the meta-analysis and FDR correction in EUR: greater CBCL lying/cheating, waking up frequently during the night, mature-rated video game frequency and weekday screentime, and spending time with peers who have been suspended and are rule-breaking, as well as lower school grades (Meta |B|≥0.036, *p*≤3.44e-05, *p_Bonferroni_*≤0.047; EUR |B|≥0.036, *p*≤4.17e-04, *p_FDR_*≤0.012, *p_Bonferroni_*≥0.71; AFR |B|≤0.072, *p*≥9.48e-03, *p_FDR_*≥0.20). Q-tests indicated that the ancestry-specific effect sizes for these phenotypes were not significantly heterogeneous (Q≤0.94*,* Q*_Pval_*≥0.33) and that percent of the total variance attributed to heterogeneity (i.e., I^2^) was 0 for all 15 effects. For the 10 phenotypes Bonferroni-significant in EUR but not significant following multiple testing correction in the meta-analysis, there was some evidence of heterogeneity in effect sizes (6 Q*_Pval_*≤0.047, 4 Q*_Pval_*=0.075-0.18; I^2^=45.7-89.0; but these should be interpreted with caution)

**References**

1.	von Hippel PT. The heterogeneity statistic I2 can be biased in small meta-analyses. *BMC Med Res Methodol*. 2015;15(1):35. doi:10.1186/s12874-015-0024-z

2.	Huedo-Medina TB, Sánchez-Meca J, Marín-Martínez F, Botella J. Assessing heterogeneity in meta-analysis: Q statistic or I2 index? *Psychol Methods*. 2006;11(2):193-206. doi:10.1037/1082-989X.11.2.193

1. The mediation package’s mediate() function outputs values of “0” for p-values that are less than 2.2e-16; for the purposes of multiple testing correction, these values were assigned a value of 2.2e-16, as also noted in Supplemental Table 35.
